# Measuring enteric pathogen force of infection through antibody responses in children

**DOI:** 10.64898/2026.09.29.26364344

**Authors:** Nikolina Walas, Lesly Simbaña Vivanco, Stuart Torres Ayala, Chabier Coleman, E. Brook Goodhew, Kelsey J. Jesser, Nicolette A. Zou, Christine S. Fagnant-Sperati, Jesse Contreras, Caitlin Hemlock, Jeffrey W. Priest, Richelle C. Charles, Edward T. Ryan, Claire Munroe, Robert L. Atmar, Julianna Colado, Hadley Burroughs, Manuel Calvopiña, William Cevallos, Josefina Coloma, Gwenyth O. Lee, Gabriel Trueba, Joseph N.S. Eisenberg, Karen Levy, Benjamin F. Arnold, the ECoMiD Investigators

## Abstract

Enteric pathogens account for a substantial global disease burden, yet population-based surveillance remains limited by the transient nature of pathogen shedding, which constrains the sensitivity of stool-based molecular testing to a narrow detection window. Detection of IgG responses in serological surveys could enable new insights into enteric pathogen transmission, but benchmarking serological measures relative to stool-based measures of infection remains a critical evidence gap. We compared measures of disease transmission in longitudinal birth cohort samples using multiplex IgG (1,575 dried blood spots, 369 children) and PCR assays (2,229 stool samples, 369 children) that overlapped for nine pathogens (norovirus GI/GII, *Shigella*/enteroinvasive *Escherichia coli* (EIEC), *Campylobacter* spp., enterotoxigenic *Escherichia coli* (LT-ETEC), *Salmonella enterica*, *Giardia* spp., *Cryptosporidium* spp., *Entamoeba histolytica*). *Campylobacter* and LT-ETEC ranked highest while *E. histolytica* and *S. enterica* ranked among the lowest by both IgG and PCR measures of infection. Measures of infection were less aligned for norovirus GI, norovirus GII, *Cryptosporidium*, *Shigella*/EIEC, and *Giardia*. Despite this, for most pathogens, and across both measures, force of infection was lowest in the urban city of Esmeraldas and substantially higher in more rural populations, with relative risks in assay measures (seroconversion rate and PCR detected prevalence) of approximately 2.0 for pathogens with the largest differences (*Shigella*/EIEC and norovirus GI). Together, our results suggest that although absolute levels of infection differed between assays, IgG and PCR captured consistent relative risk of infection across geographic strata, suggesting either method can identify high-versus low-transmission settings.

## Introduction

Enteric pathogens are a major contributor to the global burden of disease, particularly among young children in low-resource settings (1–3). Burden estimates based on clinical diarrhea and mortality likely give an incomplete view of transmission, which is central to guiding preventive interventions such as vaccines and environmental improvements. Multiplex PCR platforms have advanced our understanding of the relative burden of enteric pathogens in these settings (4,5). However, stool collection is logistically difficult and the pathogen shedding window is narrow, so enteric pathogens are rarely measured in large population-based surveys.

Serology offers a potential solution, since antibody responses persist beyond the shedding window and integrate infection over time. Multiplex IgG assays could therefore support large-scale surveillance of enteric pathogen transmission, immunity, and burden (6–9). Serological surveys have estimated that *Campylobacter* and *Salmonella enterica* incidence in Europe to be 2-6 orders of magnitude higher than passive surveillance estimates (10–13) and have found far higher incidence than previously known for *Vibrio cholerae* and *S. enterica* serovar Typhi in the Global South (14–16). Studies of children in the Global South have also shown high enteric pathogen seroconversion in the first 2-3 years of life (6,17,18). To our knowledge, no large-scale study has compared epidemiologic inference from serology with PCR detection in stool.

Before enteric pathogens can be incorporated into serological surveillance platforms, antibody-based measures of infection must be benchmarked against stool-based samples across populations. This benchmarking has been done for malaria (19,20), schistosomiasis (21), trachoma (22,23), COVID-19 (24–26), typhoid fever (15), and cholera (27), but not for the broader set of enteric pathogens responsible for most of the diarrheal disease burden worldwide.

Our objectives were to estimate enteric pathogen force of infection from seroconversion rates across an urban-rural gradient in coastal Ecuador, and to benchmark these estimates against PCR detection in stool from ages 3 to 24 months. The first two years of life are a key window of seroconversion in high-transmission settings (6,15,18). Community characteristics such as population density, access to resources, and landscape features are an important source of heterogeneity in enteric pathogen transmission (28), and stool-based testing in this cohort showed large differences in PCR-detected infections along the urban-rural gradient (29,30). Paired longitudinal blood and stool samples tested in multiplex for nine viral, bacterial, and protozoan pathogens provided a unique opportunity to compare these measures.

## Results

### Participant and sample characteristics

Our analyses include longitudinal dried blood spot and stool samples collected from the ECoMiD birth cohort in Esmeraldas Province in Northwestern Ecuador. Children were enrolled from settings along an urban-rural gradient that included the urban city of Esmeraldas, the commercial or “intermediate” town of Borbón, and rural villages accessible by road (‘rural road’) and river (‘rural river’) (**Figure 1**). Due to the small number of samples from rural river communities, we aggregated rural road and rural river into a larger rural community category for analysis. Within this cohort, 369 children contributed 1,575 blood samples (median per child 5, interquartile range 4 –5) and 2,229 stool samples (median per child 7, interquartile range 5 – 7) to the analyses (**Supplemental Table 1**). Children from the city of Esmeraldas lived in households with slightly higher levels of wealth, maternal education, crowding, cement flooring, and sanitation and lower levels of *Escherichia coli*, a fecal indicator bacterium, in their water source compared to children enrolled from more rural settings (**Supplemental Table 2**).

**Figure 1.**
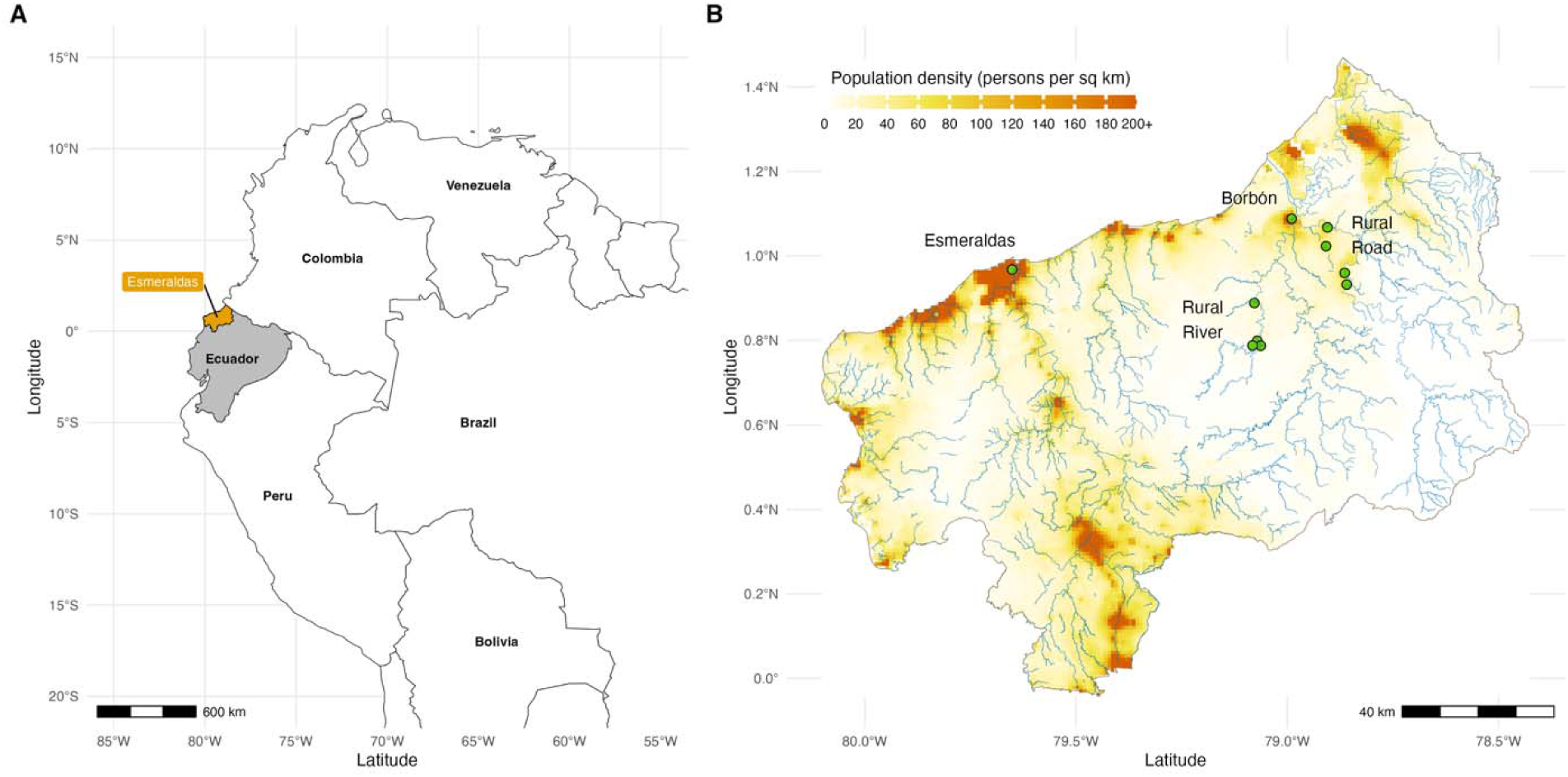
Study location along an urban-rural gradient in coastal Ecuador. **(A)** The ECoMiD birth cohort is located in Esmeraldas Province, northern coastal Ecuador. **(B)** The study included mothers and their newborn children from communities along an urban-rural gradient, including the city of Esmeraldas (population approximately 162,000), the town of Borbón (population approximately 5,000), four road-accessible rural villages (‘rural road’; populations 500 to 1000) and four river-accessible rural villages (‘rural river’; populations 200 to 700). Major rivers (blue) were provided by OpenStreetMap. Population density of Esmeraldas Province was provided by WorldPop.

Blood samples were tested using a multiplex bead assay on the Luminex platform for bound IgG and stool samples were tested using two multiplex PCR assays previously determined to provide comparable results (31), (Luminex xTAG-GPP at ages 3, 9, 15, 24 months and a custom TaqMan Array Card at ages 6, 12, 18 months). In this analysis, we focused on nine pathogens measured in both assays: norovirus GI and GII, *Shigella*/enteroinvasive *E. coli* (EIEC), *Campylobacter*, enterotoxigenic *Escherichia coli* (LT-ETEC), *Salmonella enterica*, *Giardia*, *Cryptosporidium*, and *Entamoeba histolytica.* We analyzed norovirus genogroups I and II separately due to large differences in their burden and force of infection, with GII accounting for 8% (PCR) and 51% (IgG) of norovirus infections in the cohort (30) and up to 89% of norovirus outbreaks globally (32–34). At the sample level, IgG responses were strongly correlated between antigens to the same pathogen, but between-pathogen correlation was modest (**Supplemental Figure 1**).

### Serology integrates infection histories over time

High levels of enteric pathogen infection were detected during the first 24 months of life. Consistent with a durable adaptive immune response, age-specific IgG point seroprevalence rose earlier and to a higher level over time compared with PCR detected point prevalence for most pathogens (**Figure 2**). In contrast, PCR detected cumulative incidence mapped closely to IgG point seroprevalence with the exception of norovirus GI/GII and *Giardia.* Because cumulative incidence does not decline once accrued, it is unaffected by antibody waning or boosting and consistently exceeded point prevalence, with the largest gaps for IgG evident in norovirus GII, *Giardia*, and *Cryptosporidium.* By 24 months, IgG seroprevalence was highest for LT-ETEC (82%, 95% CI 77% to 87%), norovirus GII (84%, 95% CI 80% to 88%) and *Campylobacter* (78%, 95% CI 73% to 83%) and *Shigella*/EIEC (71%, 95% CI 66% to 76%). PCR detected prevalence remained constant or plateaued after 9 months for most pathogens, with the exception of *Giardia* and *Shigella*/EIEC. Interestingly, PCR detected prevalence of *S. enterica* decreased over the study period. IgG seroprevalence was generally higher over the study period for all pathogens, except *Giardia*, in Borbón and rural communities compared to the city of Esmeraldas (**Figure 3**). Over the course of the study, *Campylobacter* had the highest overall prevalence by IgG and LT-ETEC had the highest overall prevalence by PCR (**Supplemental Table 3**).

**Figure 2.**
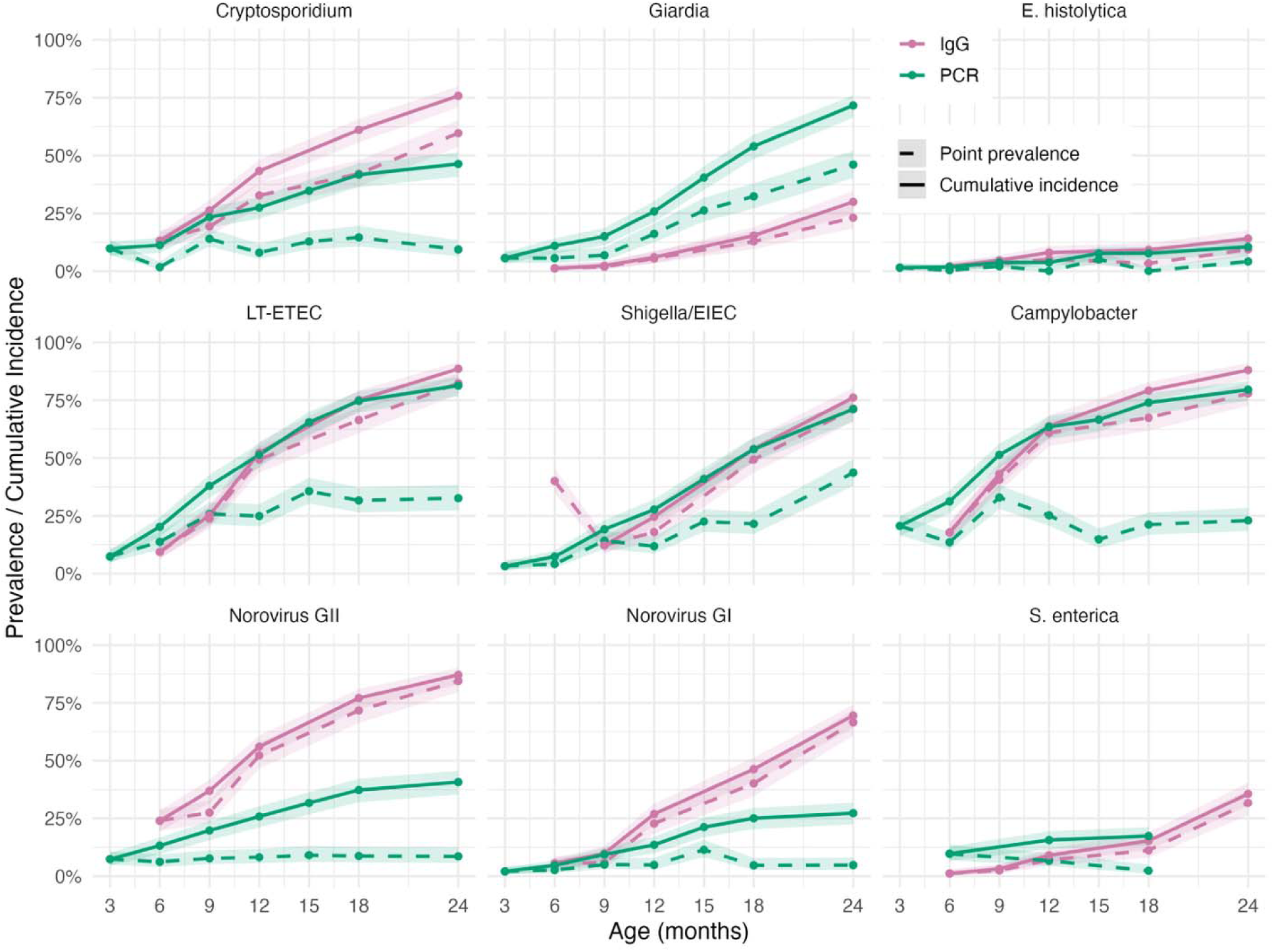
Point prevalence and Kaplan-Meier estimated cumulative incidence of first pathogen detection assayed by both PCR and IgG serology in the ECoMiD cohort. Prevalence (positives per number tested each round) and cumulative incidence (1 − Kaplan-Meier survival estimate, Greenwood 95% CI) at each age are shown by assay, with shaded 95% confidence bands. Cumulative incidence lines begin at each pathogen and assay’s first observed testing round rather than a common start, reflecting staggered entry into testing. The 6-month *Shigella*/EIEC IgG reading was excluded from the cumulative incidence estimate (not point prevalence) to account for probable maternal antibody contribution.

**Figure 3.**
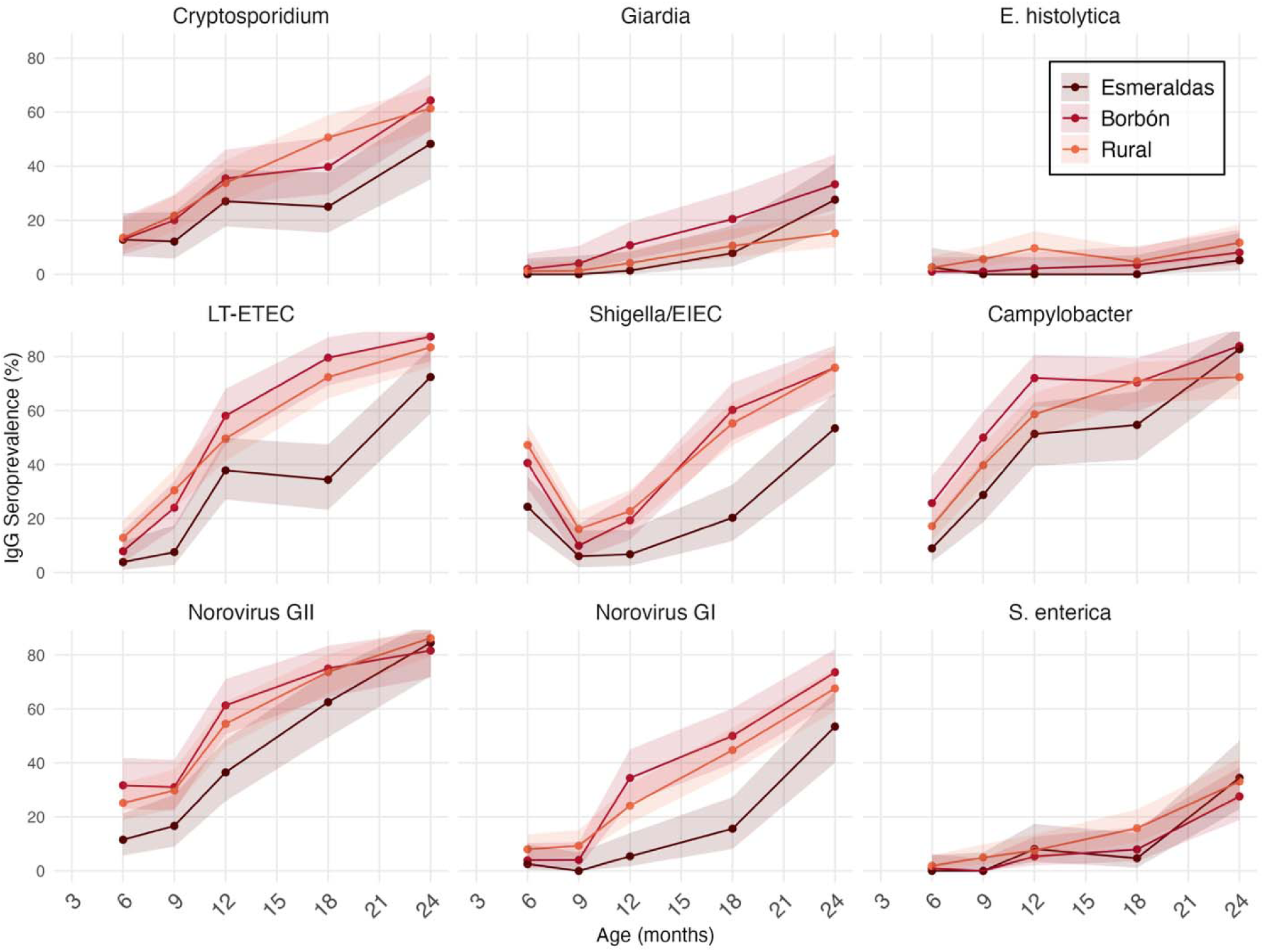
IgG seroprevalence by age and community type across an urban-rural gradient in coastal Ecuador. IgG seroprevalence at each age estimated from children enrolled in the cohort (city of Esmeraldas, N=84; Borbón, N=108; Rural, N=177). IgG serology detects the IgG antibody levels from dried blood spots using a multiplex bead assay. Shaded bands represent 95^th^ confidence intervals around the mean.

Longitudinal IgG measurements captured complex infection dynamics over the study period. For example, maternal IgG contributions in marginal age-seroprevalence curves were only evident for *Shigella/*EIEC with a decline from ages 6-9 months (**Figure 2**), but individual child trajectories illustrated IgG waning through 9 months was common for many other pathogens beyond *Shigella*/EIEC (**Supplemental Figures 2, 3, 4**). Age-stratified estimates of the seroconversion rate (SCR) and seroreversion rate (SRR) showed a predominance of seroreversion in the first year of life and a consistent increase in the SCR after 12 months, a period that coincides with increased mobility and introduction of complementary foods (**Supplemental Figure 5**). Higher SRRs from 6-9 months likely reflect a combination of maternal IgG waning and less robust immune responses in infancy.

Analyses of PCR-detected infections in the cohort focused on prevalence and cumulative incidence because the short duration of infection relative to the three-month sampling intervals meant we would substantially underestimate incidence rates. Indeed, for all pathogens studied, a substantial number of children seroconverted who were not detected positive by PCR during the quarterly stool testing (**Supplemental Figures 2, 3, 4**). Most children who tested positive by PCR also seroconverted, illustrating how IgG provides a more comprehensive picture of recent infection history. An exception was *Giardia,* where 106 children tested PCR positive before the last recorded serology result but did not seroconvert. The subgroup that failed to seroconvert tended to have quantitative PCR values closer to positivity cutoffs than those who seroconverted, though this finding was less prominent in rural sites. This may represent low-intensity infections, potential ingestion of inactivated oocysts that failed to stimulate an adaptive immune response, or transient colonization with animal-infecting *Giardia* strains (**Supplemental Figure 6**).

### Within- and between-pathogen consistency in epidemiologic estimates

PCR and serology measure different dimensions of the infection and immunity process, yet there was a strong relationship between measures from the two assays across pathogens and community settings (**Figure 4**). Both PCR detected prevalence and PCR detected cumulative incidence by 24 months aligned well with estimated SCR (Spearman ρ=0.81, 95% CI: 0.70, 0.86 and ρ=0.81, 95% CI: 0.66, 0.84, respectively). *Giardia* was an outlier in the comparison, where the IgG seroprevalence was far below what might be expected given stool-based infection (**Figure 2**) which could be explained by a large number of children who had low-intensity PCR-detected infections but failed to seroconvert (**Supplemental Figure 6**). Additionally, the SCR for norovirus GI and GII appeared to be proportionally higher compared with PCR detected prevalence in relation to the generally linear relationship across pathogens and community types, potentially due to the moderate sensitivity of multiplex PCR assays for norovirus (78-88% sensitivity (35)) or a short viral infection and shedding duration that may not be captured during the PCR testing window (**Supplemental Figure 9**).

**Figure 4.**
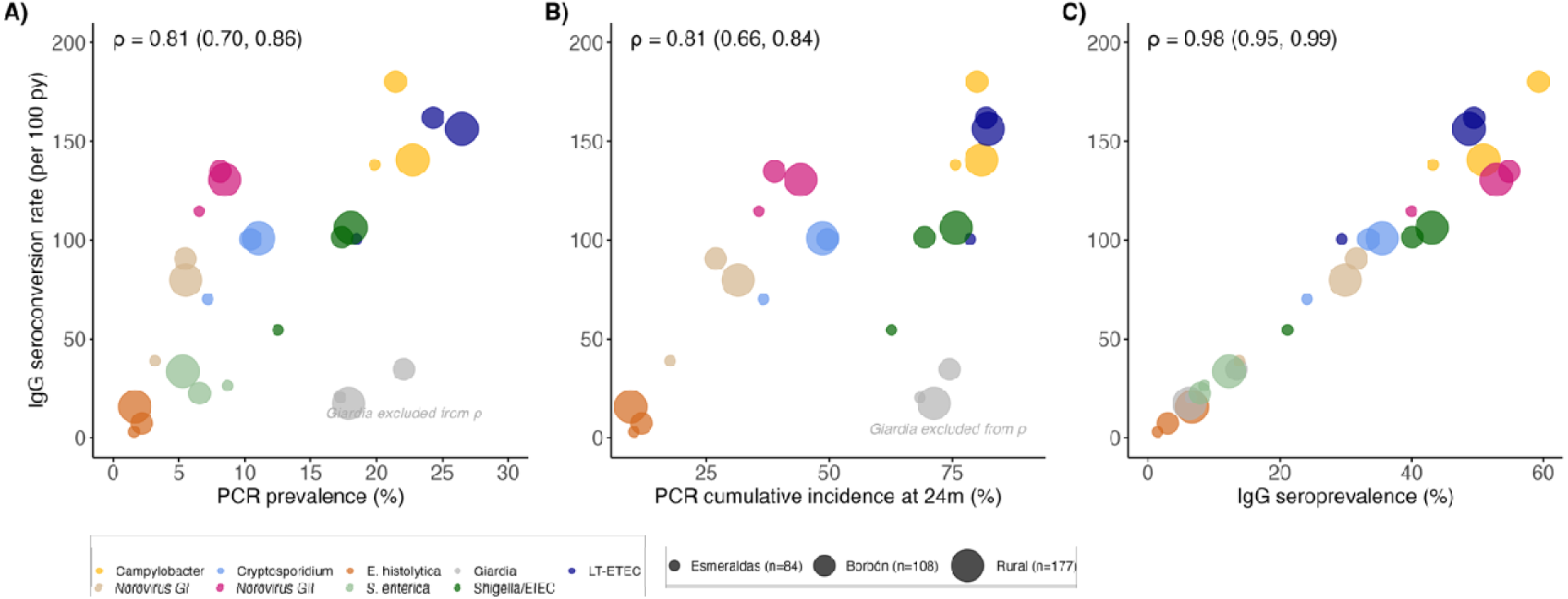
Relationship between measures of pathogen detection by PCR and serology, across pathogens and community types (Esmeraldas, Borbón, Rural). IgG seroconversion rate and IgG seroprevalence were estimated over five visits from ages 6 to 24 months (n=369 children, 1575 samples). PCR prevalence was estimated over seven visits from ages 3 to 24 months (n=369 children, 2229 samples). PCR cumulative incidence at 24 months was estimated using the Kaplan-Meier estimator to account for left-truncated entry and loss to follow-up. Point size reflects the number of children tested in each community type. Spearman correlation coefficients (ρ) were calculated directly from the observed pathogen-community estimates. 95% confidence intervals for ρ were estimated using nonparametric bootstrap, resampling children with replacement. *Giardia* was excluded from the Spearman correlation estimate (ρ) in panels A and B because the relationship between measures differs markedly from other pathogens in this setting (details in Results). (A) Incident seroconversion rate compared with PCR prevalence (with *Giardia* included ρ = 0.63, 95% CI: 0.51, 0.67). (B) IgG seroconversion rate compared with PCR-based cumulative incidence at 24 months (with *Giardia* included ρ = 0.66, 95% CI: 0.49, 0.68). *S. enterica* PCR results were not available at 24 months and were excluded (C) Incident seroconversion rate compared with IgG seroprevalence across all visits.

Despite complex IgG dynamics over the age range, overall seroprevalence and the SCR were almost perfectly linearly related across pathogens and community types (**Figure 4C**; Spearman ρ=0.98, 95% CI: 0.95, 0.99). Since seroprevalence and the SCR were estimated non-parametrically this strong relationship between summary measures is likely not an artifact of statistical modeling. SCR and SRR aligned linearly, though the relationship was less clear (Spearman ρ=-0.79, 95% CI: -0.84, -0.52), likely due to the small number of seroreversions given the limited study time window **(Supplemental Figure 5 and 7)**. This correlative relationship was maintained across community types, though SCR and PCR-detected prevalence (0.93, 95% CI: 0.78-0.93) and SCR and SRR (-0.85, 95% CI: -0.92, -0.68) were more strongly correlated in rural communities (**Supplemental Figure 8**). This agreement is supported by a similar rank detection by each assay. *S. enterica* and *E. histolytica* fell exactly on the identity line; *Campylobacter*, LT-ETEC, *Cryptosporidium* and *Shigella*/EIEC differed by one rank; norovirus GI, norovirus GII and *Giardia* differed by two, three and five ranks. This pattern was consistent across community types: *Giardia* was ranked substantially higher by PCR than by IgG serology in Esmeraldas, Borbón, and rural communities alike, while norovirus GII showed the inverse, ranking higher by IgG than by PCR in all three settings (**Supplemental Figure 9; Supplemental Table 4**).

### Heterogeneity in transmission along the urban-rural gradient

A key rationale for IgG testing in the cohort was a previous finding of large differences in pathogen infection between children in the city of Esmeraldas, Borbón, and rural communities using the TAC PCR assay (29,30). We anticipated that serological measures of transmission would provide similar inference to PCR in terms of relative changes in population-level measures of infection along the urban-rural gradient. Relative changes in PCR detected prevalence and the SCR along the urban-rural gradient were broadly consistent with one-another across pathogens, with higher levels of infection and seroconversion in Borbón and rural communities compared to the city of Esmeraldas (**Figure 5**). Our primary focus was incident seroconversion, but we additionally estimated stratified age-seroprevalence curves by community type (**Figure 3**). Age-seroprevalence curves that rose more slowly in the city of Esmeraldas were consistent with lower force of infection compared with Borbón and rural communities. Pathogens and settings with the highest force of infection as measured by the SCR also had the lowest SRR, likely from continuous boosting of IgG due to repeated infections (**Supplemental Figure 7**). Despite capturing distinct aspects of transmission—current detection by PCR versus cumulative exposure with IgG — both approaches yielded consistent epidemiological inferences regarding relative transmission intensity along the urban-rural gradient.

**Figure 5.**
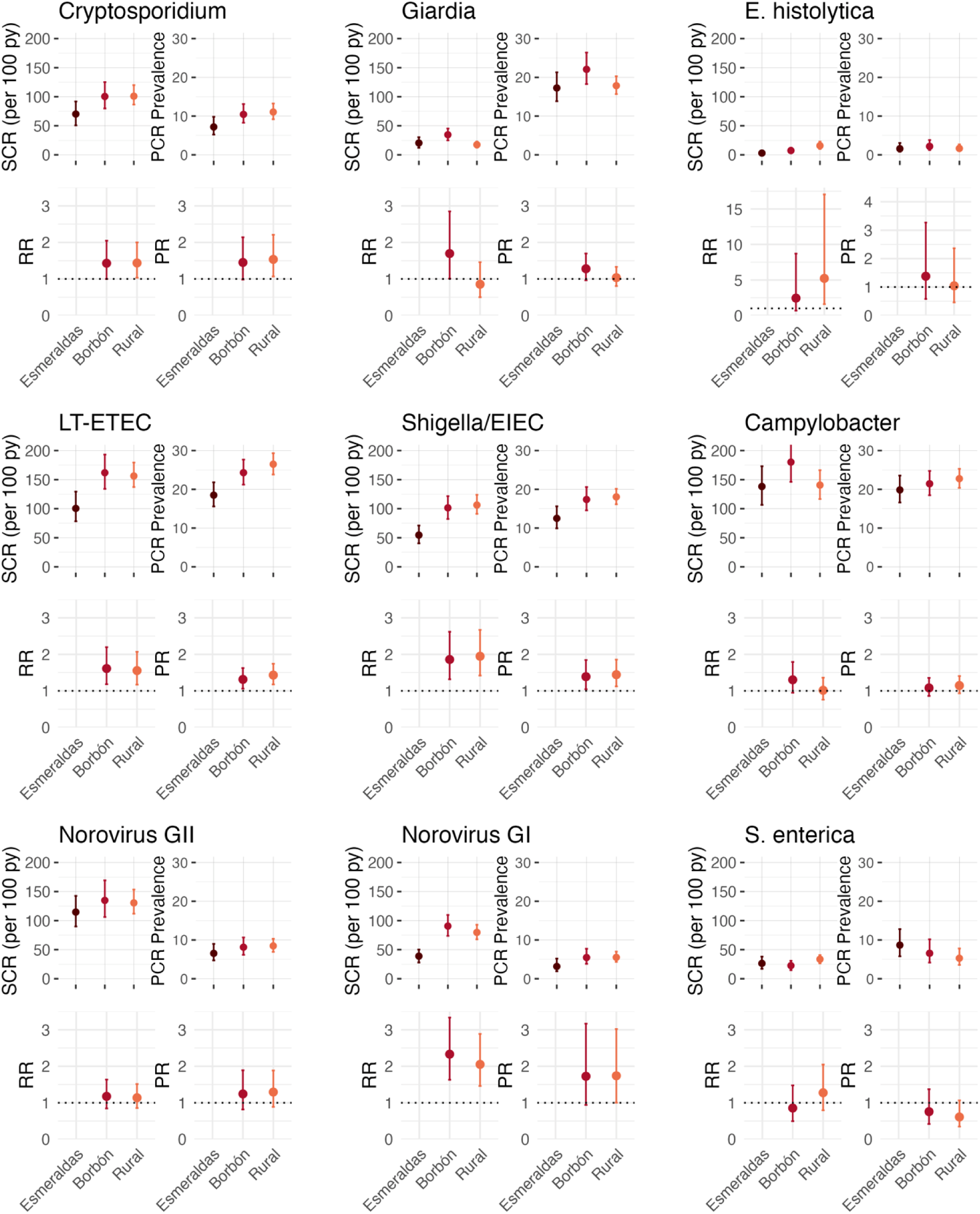
Pathogen-specific measures of relative risk along an urban-rural gradient assessed by the seroconversion rate and PCR prevalence. Seroconversion rates (SCR per 100 person-years) and PCR-based infection prevalence in stool represent averages through age 24 months by three community types—city of Esmeraldas, Borbón, and Rural villages. Rate Ratios (RR) for the SCR and Prevalence Ratios (PR) for infection use the city of Esmeraldas as the reference group. Error bars represent 95% confidence intervals.

## Discussion

In this large-scale comparison of serological and stool-based measures of enteric pathogen infection, we found that the two approaches yielded consistent measures of relative risk across community types. For most pathogens, children in the city of Esmeraldas experienced lower seroconversion and infection in their first two years of life than children in more rural settings (Figure 5). Moreover, the rank-ordering of infection across communities and across pathogens was largely preserved between assays (Figure 4; Supplemental Figure 9). Where the two approaches diverged was in the absolute level of infection detected: IgG-based measures revealed higher cumulative burden than would be inferred from periodic stool collection alone, reflecting its integration of exposure over age compared with active intermittent pathogen shedding.

We found the relative risk of seroconversion and PCR detected infection was higher in rural communities and Borbón compared to the city of Esmeraldas for *Cryptosporidium*, *Shigella*/EIEC, LT-ETEC, and norovirus GI using both PCR and serological measures. In contrast, there was no consistent increase in risk along the urban-rural gradient for *Giardia, Campylobacter*, norovirus GII, or *S. enterica*. The consistency between assays in this absence of heterogeneity along the urban-rural gradient was still informative and was reflected in consistent, rank-preserving order between seroconversion rates and PCR detected prevalence across all pathogens and settings.

An outlier in the analysis was *Giardia,* which showed consistent measures of relative risk between serological and stool assays along the urban-rural gradient (Figure 5), but had far lower IgG seroconversion rates and IgG seroprevalence relative to PCR detected prevalence in context with other pathogens (Figure 2, Figure 4). There are a few potential explanations for the discrepant results. Firstly, unlike many invasive pathogens, *Giardia* remains confined to the intestinal lumen and does not invade host tissues. Consequently, protective immunity is largely mediated by local mucosal responses, particularly secretory IgA, rather than systemic IgG. Despite this, IgG responses have been observed in other communities (36–38). Secondly, young children may have immature anti-*Giardia* humoral responses. Several studies suggest that effective antibody responses against *Giardia* improve with age and repeated exposure (39); however, we did not observe a similar age-related trend for other pathogens with comparable exposure dynamics. Lastly, the PCR target used here (18S) does not distinguish between human- and non-human–infecting *Giardia* assemblages. Given reports of humans harboring animal-associated *Giardia* assemblages in the region, especially assemblage D prevalent in dogs, it is possible that children are carrying non-infectious organisms detectable by PCR without eliciting a detectable host IgG response (40–42). Additionally, *Giardia* has been shown to behave as a commensal organism, permitting low-level shedding without eliciting immune activation (36,43). This interpretation is further supported by our finding that many PCR-positive children who did not seroconvert had MFI and Ct values just above the positivity threshold, which may also reflect very low-intensity infections insufficient to mount a strong immune response. Although these hypotheses remain speculative, this discrepant finding for *Giardia* motivates further comparisons between serological and stool-based measures of infection in other populations to determine whether this pattern is specific to coastal Ecuador or reflects a broader trend.

For many pathogens, seroprevalence steadily increased after nine months, likely due to incident boosting from ongoing exposure as immunity to most enteric pathogens is not long-lived (44). Differences between serological and stool-based measures were most stark for norovirus GI and GII and *Cryptosporidium*, where PCR detected prevalence remained low (∼10%) across ages while IgG seroprevalence consistently increased to 60-84% by age 24 months (Figure 2). For norovirus, this discrepancy between infection prevalence and seroprevalence may reflect a short duration of five to ten days for viral shedding or lower viral loads of asymptomatic infections, which are common in high-transmission settings but still induce IgG responses (45–47). Alternatively, multiplex PCR assays, such as the GPP assay, may have low sensitivity for viral targets compared with optimized singleplex assays, particularly at low viral concentrations (48,49). These three pathogens also had the highest seroconversion rates relative to PCR-detected prevalence (Figure 4). Individual IgG trajectories showed repeated boosting and waning, which suggests that the seroconversion rates captured reinfections (Supplemental Figures 2–4). Together, these findings suggest that the relationship between PCR and serological measures varies slightly across pathogens and is influenced by the underlying infection dynamics.

Longitudinal sampling enabled us to compare incident seroconversion rates with seroprevalence. Although repeated seroconversion and seroreversion was common for many pathogens (Supplementary Figure 2, 3, 4, 5, 7), an interesting result was that longitudinal seroconversion rates were linearly related with seroprevalence across pathogens and community types (Figure 4). The empirical, linear relationship between longitudinal seroconversion rates and seroprevalence suggests that estimating seroprevalence in cross-sectional surveys may be sufficient to summarize differences in transmission even in light of highly complex IgG dynamics over the age range. This is useful because many large-scale public health serosurveys are cross-sectional, and it suggests that longitudinal cohorts would not be essential to characterize heterogeneity in transmission.

Our findings should be interpreted with considerations to generalizability. We restricted serological measurements to children aged 6–24 months, which limits inference for older age groups. However, this window captures the period of most rapid seroconversion in high-transmission settings, where early and intense exposure drives IgG production. Measurements at older ages may yield less information given near-universal seroprevalence, and IgG interpretation in adults is further complicated by antibody waning. For pathogens with lower transmission intensity, such as *S. enterica* serovar Typhi or *V. cholerae*, methods that estimate time-since-infection may offer additional insight, though whether these approaches are applicable to the high-transmission enteric pathogens studied here remains unclear. Our study was also conducted in a single region of Ecuador, but the communities we studied span multiple populations, with a wide range of transmission intensities across pathogens and community types. Replication in other populations would be valuable as the divergent *Giardia* results noted above suggest that local epidemiology may influence the relationship between the two assay measures. Seroconversion rates and seroprevalence were broadly lower than those reported in similar age groups (6), though the rank order of pathogen prevalence was consistent. This observation suggests that, while absolute measures may be community-specific, broader pathogen patterns may be generalizable.

Our analysis compared overall measures of infection risk along the urban-rural gradient using IgG and PCR and was not designed to identify underlying causes. Nevertheless, the higher enteric pathogen risk observed in Borbón and rural villages relative to the city of Esmeraldas is consistent with documented declining infrastructure and increased environmental exposures across this gradient (Supplemental Table 2), factors that influence both pathogen persistence and transmission intensity. For example, inadequate access to improved water and sanitation in rural settings may sustain transmission of enteric pathogens (50,51). Borbón may also serve as a transmission hub, seeding infection to surrounding rural villages via river and road networks (52).

This study had limitations. First, at the individual level we observed a high correlation between antigens from the same pathogen (Supplemental Figure 1). *S. enterica* lipopolysaccharide (LPS) Groups B and D antigens share antigenic determinants within their LPS structures (53), *Campylobacter* antigens p18 and p39 are highly conserved across pathogenic species (54) and structural similarities between norovirus genogroups are all likely driving broadly reacting IgG responses (55,56). This suggests that seroepidemiologic analyses for these pathogens may lack species specificity compared to molecular based assays such as PCR.

A second limitation concerns the derivation of seropositivity thresholds. We were unable to curate sample sets of confirmed positive and negative sera to derive seropositivity thresholds for many enteric pathogens, given how common they are in populations around the world. Our alternative approach of constructing a pre-exposed endemic cohort to estimate thresholds aligned well with receiver operating characteristic thresholds, but it requires longitudinal sampling. The absence of off-the-shelf serological assays and well-characterized positive control sera for enteric pathogens limits both the potential for widespread adoption and the standardized establishment of cutoff values.

Our PCR results also had limitations. We used two separate assays for PCR testing, one of which (GPP) did not provide target-level data. We have demonstrated alignment in PCR results for the pathogens included here and excluded GPP data for *S. enterica* at ages 3, 9, 15 and 24 as it was not comparable between the two assays (57). Additionally, PCR testing was periodic and may not have captured the full infection history for each child, though this study represented the most intensive enteric pathogen surveillance available.

We have shown that antibody-based measures complement stool-based PCR testing because antibody responses remain elevated for many months after infection, providing more information in studies with infrequent measurements. The close alignment between seroprevalence and seroconversion rates further suggests that cross-sectional surveys may be sufficient to characterize heterogeneity in enteric pathogen transmission, without the resource demands of longitudinal cohort studies. Longitudinal serological data provide an additional opportunity to characterize within-host infection dynamics — including boosting, waning, and reinfection patterns — that cannot be captured by prevalence estimates alone. The interpretation of serological measures will need to account for local epidemiological context for pathogens where discordance due to infection dynamics is most likely.

### Conclusions

In a setting with large variation in enteric pathogen risk, serological and PCR based measures of transmission provide consistent inference of population-level differences in transmission. Our findings suggest that seroconversion rates provide a sensitive and dynamic measure of transmission intensity, while remaining broadly consistent with seroprevalence in high-transmission settings with frequent antibody boosting and waning. These results support the use of population-based, serological surveys in young children to assess enteric pathogen transmission and inform public health programs.

## Material and Methods

### Study population and setting

We compared molecular and serological estimates of enteric pathogens detected in children enrolled in the ECoMiD (‘*Enteropatógenos, Crecimiento, Microbioma, y Diarrea’*) prospective birth cohort that enrolled mothers and their newborn children during prenatal visits in Esmeraldas Province, Ecuador (58). The study enrolled 521 children from May 15, 2019 to October 29, 2022, from ten communities across an urban-rural gradient (Figure 1). IgG testing focused on a sub-cohort of 369 children that were enrolled after the SARS-CoV-2 shutdown. Study communities were classified into three groups (i) The city of Esmeraldas, the provincial capital with an approximate population of 162,000; (ii) Borbón, a town serving as the commercial hub of a designated rural region, with about 5,000 residents (intermediate site); (iii) road-accessible rural villages surrounding Borbón (Timbiré, Selva Alegre, Colon Eloy, and Maldonado), each with populations ranging from roughly 500 to 1,000 (rural road villages); and (iv) more remote rural villages (Santo Domingo, Zancudo, Colon de Onzole, and San Francisco) which are primarily reachable by river or have limited road access and populations of approximately 200 to 700 (rural river villages). Due to the small number of children from rural-river communities, analyses combined rural river and rural road communities into a single “rural” community group. Sample processing and laboratory methods are described in detail in supporting information.

### Statistical analysis

#### Determination of seropositivity thresholds

The serology multiplex assay generates quantitative measures of bound IgG. From continuous IgG distributions we determined seropositivity thresholds to enable estimates of seroprevalence and seroconversion rates. Seropositivity thresholds for *Cryptosporidium* (Cp17, Cp23), *E. histolytica* (LecA), and *Giardia* (VSP-5) antigens were determined using a receiver operator characteristic curve analysis of known positive and negative samples at the US Centers for Disease Control and Prevention (CDC) (59,60). Seropositivity thresholds for the remaining antigens were determined using a pre-exposed endemic cohort approach (44,60). For each antigen, we identified children who had a 100-fold increase in MFI-bg response between two visits, limited to the child’s first boost in the case of multiple, large increases during follow-up. The IgG measurement that preceded the 100-fold increase was assumed to represent an unexposed value from which we estimated seropositivity thresholds as the mean plus three standard deviations. We required a large, 100-fold increase in IgG as an empirical decision to prevent the inclusion of baseline IgG levels that might be mid-boost following an infection, which could lead to over-estimating the variability in the unexposed distribution. Thresholds were comparable for antigens with values estimated by both methods (**Supplemental Figure 10**).

#### Prevalence estimates

We estimated point seroprevalence and PCR-detected prevalence as the proportion of samples positive divided by the number measured at each age. We estimated overall seroprevalence and PCR-detected prevalence as the proportion of samples positive over the study period by the number of samples measures over the study period. To estimate 95% confidence intervals, we used an intercept-only logistic regression model with robust standard errors clustered at the child level to account for repeated measures within children. For age specific estimates, where each child contributes one sample, we used Wilson score intervals. We estimated cumulative incidence of first pathogen detection (PCR or IgG seropositivity) using the Kaplan-Meier estimator, fit separately by pathogen and stratified by assay. Children were left-truncated at their first observed testing round using counting-process notation in the survival package (R), with an internal missing round carried forward as the child’s prior status and a run of missing rounds with no prior positive treated as right-censoring at the first missed round. Children with no trailing missingness were administratively censored at their last observed round. A child positive at their first observed round was counted as an event at that round. The 6-month Shigella/EIEC IgG reading was excluded to account for probable maternal antibody contribution. We used Greenwood’s formula to estimate the variance of the Kaplan-Meier estimator and constructed 95% confidence intervals on the log-transformed scale, as implemented in the survival package in R.

#### Force of infection from prospective seroconversion rates

The SCR is one measure of a pathogen’s force of infection (61,62). We defined incident seroconversion as a change from seronegative to seropositive between visits and assumed events occurred at the midpoint between visit dates to estimate person-time at risk. Intervals for which person-time could not be calculated because of a missing visit date were excluded (one seroconversion). We estimated seroconversion rates as events per 100 person-years overall and stratified by community type and by age (< 12 months vs ≥ 12 months). We estimated SRRs similarly, but defined seroreversion as a change from seropositive to seronegative. We estimated 95% confidence intervals for rates using a nonparametric bootstrap that resampled children with replacement to account for within-child correlation (1,000 replicates).

#### Quantifying differences in infection along the urban-rural gradient

We compared pathogen-specific PCR detected prevalence by community type using children in the city of Esmeraldas as a reference group and estimated the relative changes in prevalence for Borbón and rural communities using a modified Poisson regression with a log link and indicator variables for the other community types, using robust standard errors clustered at the child level (63). Similarly, we estimated relative changes in the SCR using Poisson regression models with indicators for community type, person-time as an offset, and robust standard errors clustered at the child level. We exponentiated coefficients from each model to estimate prevalence ratios (PR) for PCR analyses and rate ratios (RR) for serology analyses.

#### Epidemiologic comparisons of serology with stool-based measures of infection

We assessed consistency between serological and stool-based (PCR) measures of infection by computing Spearman rank correlations across pathogens for PCR detected prevalence, PCR cumulative incidence at 24 months, seroprevalence, SCR, and SRR, both pooled across the full cohort and separately within each community type (Esmeraldas, Borbón, and Rural) to evaluate whether this concordance was consistent across settings. We used a nonparametric bootstrap, resampling children with replacement, to estimate 95% confidence intervals for the Spearman correlation coefficients, bounded by the 2.5th and 97.5th percentiles of the bootstrap replicate distribution (2,000 replicates). For community-stratified correlations, children were resampled separately within each community type at that community’s own sample size (Esmeraldas, n = 84; Borbón, n = 108; Rural, n = 177). To assess within-pathogen consistency in epidemiologic summaries of serological and stool-based measures of infection, we compared relative changes in prevalence and the SCR along the urban-rural gradient.

### Artificial intelligence disclosure

The authors used Claude 5.0 Sonnet (Anthropic) for code generation, processing and cleaning, and for non-generative writing assistance. All output was reviewed by the authors, who take full responsibility for the content of the manuscript.

### Ethics

The study protocol was reviewed and approved by institutional review boards at the University of Washington (UW; STUDY00014270), Emory University (IRB00101202), the University of California, San Francisco (21–33932), Universidad San Francisco de Quito (USFQ; 2018-022M), and the Ecuadorian Ministry of Health (MSPCURI000253-4). Prior to sample collection, field staff obtained written informed consent from each child’s guardian. Recruitment took place between May 15, 2019, and October 29, 2022.

## Data Availability

Due to the small population size of the study region, our institution's IRB determined that individual-level data, even after de-identification, could be considered potentially identifiable given the constraints of participant traceability. All code used in analysis to display the figures are available on Zenodo (DOI: 10.5281/zenodo.23376476). Inquiries for access to individual-level data underlying this analysis can be directed to the corresponding author.

## Author contributions

Following CRediT taxonomy, conceptualization (NW, BFA), data curation (STA, LSA, KJJ, NZ, JContreras, JColado, CSF, NW), formal analysis (NW, BFA), funding acquisition (GT, JNSE, KL, BFA), investigation (NW, LSV, STA, CC, EBG, HB), methodology (NW, BFA, RCC, ETR), project administration (NW, LSV, STA, CC, WC, HB, GOL, GT, JNSE, KL, BFA), software (NW, JC, BFA), supervision (GOL, GT, JNSE, KL, BFA), validation (NW, CC, EBG), visualization (NW, BFA), Writing—Original Draft Preparation (NW, BFA), Writing—Review & Editing (LSV, STA, NW, CC, EBG, KJJ, NZ, CSF, JContreras, CH, JP, RCC, ETR, CM, RA, JColado, HB, MC, WC, JC, GOL, GT, JNSE, KL, BFA)

## Acknowledgements

The authors thank Gretchen Cooley and Diana L. Martin at the US Centers for Disease Control and Prevention for their multiplex serology antigen bead coupling, protocols, laboratory training and ongoing support. The authors would like to acknowledge Dr. Jan Vinjé at the US Centers for Disease Control and Prevention for his contributions to the development of serological assays for norovirus antigens. The authors thank the participating families for enrolling in the ECoMiD study and providing specimens. We also gratefully acknowledge the following field staff, without whose work administering surveys and collecting samples this study would not have been possible: Diana Barreiro, Mariuxi Caicedo, Grace Macias, Maria Nela Obando, Karen Caicedo, Yesica Perlaza, Yina Segura, Jorge Mejia, Juleisy Palma, Ana Rosero, Tamara Valencia, Zaida Jimenez, Sara Charcopa, Daasny Monrroy, Denise Angulo, Naira Martinez, and Dayanara Wila. The authors thank William Petri (UVa) and Joel Herbein (TechLab) for the generous contribution of LecA protein.

## Disclaimer

Use of trade names is for identification only and does not imply endorsement by the Public Health Service or by the U.S. Department of Health and Human Services. The findings and conclusions in this report are those of the authors and do not necessarily represent the official position of the Centers for Disease Control and Prevention.

## Data availability

Due to the small population size of the study region, our institution’s IRB determined that individual-level data, even after de-identification, could be considered potentially identifiable given the constraints of participant traceability. All code used in analysis to display the figures are available on Zenodo (<u>DOI: 10.5281/zenodo.23376476</u>). Inquiries for access to individual-level data underlying this analysis can be directed to the corresponding author.

