## Supplementary material for "Measuring enteric pathogen force of infection through antibody responses in children": ecomid_medrxiv_v2_SI.docx

**This PDF file includes:**

Supporting text

Figures S1 to S10

Tables S1 to S6

SI References

Supporting Text

**Supplemental Methods**

### Stool collection and testing

Caregivers collected stool samples every three months from ages 3 to 24 months in a small, insulated container. Samples were aliquoted within 3 hours of production if refrigerated, or 1 hour of production if unrefrigerated. Aliquots were stored in liquid nitrogen for up to one month until transport to the Universidad San Francisco de Quito for long-term storage at −80 °C. Qiagen QIAamp Fast DNA Stool Mini Kit (QIAGEN, Germantown, MD) with modifications was used to extract nucleic acids from stool samples (180–220 mg) that were stored at −80 °C. Further details can be found in (1).

The study protocol included PCR testing using two multiplex assays at different six-month intervals: Luminex Gastrointestinal Pathogen Panel (xTAG GPP) at ages 3, 9, 15, and 24 months tested at the Universidad San Francisco de Quito and custom TaqMan array cards (TAC) at ages 6, 12, and 18 months tested at the University of Washington. A previous validation study demonstrated high comparability between the GPP and TAC assays in the cohort except for *S. enterica,* where the xTAG GPP assay had poor specificity in community-based samples (2) — GPP results for that target were discarded in the present analysis. The Luminex xTAG GPP protocol was used to amplify and hybridize nucleic acid samples following manufacturer’s instructions (Luminex Corporation, Austin, TX). Luminex xTAG PCR targets and sequences are proprietary and not available. xTAG RNAse-free water was used as a negative control and three stocks of known pathogen DNA (ZeptoMetrix NATtrol GI Verification Panel 2) were used as positive controls. Enteric pathogen PCR targets can be found in Supplemental Table 2. Details regarding primer sequences can be found in this original publication (3,4). Pathogen gene targets using TAC (ThermoFisher Scientific, Waltham, MA) were cycled on a QuantStudio 7 Flex instrument (ThermoFisher Scientific). Samples with cycle threshold (Ct) value <35 for any of the gene targets for a pathogen were classified as positive as described in other studies (5–7).

### Blood collection and testing

Finger prick capillary blood was collected by field staff on TropBio filter paper (Cellabs, Brookvale, NSW, Australia) with six 10 μL extensions in household visits when children were ages 6, 9, 12, 18, and 24 months. We did not collect finger prick blood samples at 3 months alongside stool samples because it was a slightly more invasive measurement at that young age and we reasoned that maternal IgG would dominate the measurement. Dried blood spot samples were stored in plastic bags with desiccant at -20^o^C until testing. Single 10 μL extensions from each sample were eluted into a buffer matrix (1X PBS pH 7.2-7.4, 0.5% casein, 0.5% PVA, 0.8% PVP, 0.3% Tween 20, 0.02% NaN3, 3 µg/ml of *E. coli* extract) overnight at 4^o^C and further diluted to a concentration of 1:400 (8). The eluate was first incubated for 1.5 hours with microsphere beads coupled to 45 antigens developed and produced by the US Centers for Disease Control and Prevention (CDC) representing a range of pathogens including enteric pathogens, neglected tropical diseases, vaccine preventable diseases, and arboviruses (9). Enteric pathogen antigen targets are outlined in detail in Supplemental Table 5. This was followed by a 45-minute incubation with biotinylated mouse anti-human IgG and IgG4 (Southern Biotech, Birmingham, AL, USA), and finally a 30-minute incubation with streptavidin conjugated to R-phycoerythrin, which served as the fluorescent marker (Invitrogen, Waltham, MA, USA). After incubation, plates were triple washed and suspended in 100 µl PBS. After storage at 4^o^C, bound IgG was measured and reported as a median fluorescence intensity *minus* background (MFI-bg) using a MAGPIX instrument and the Luminex platform.

Figures

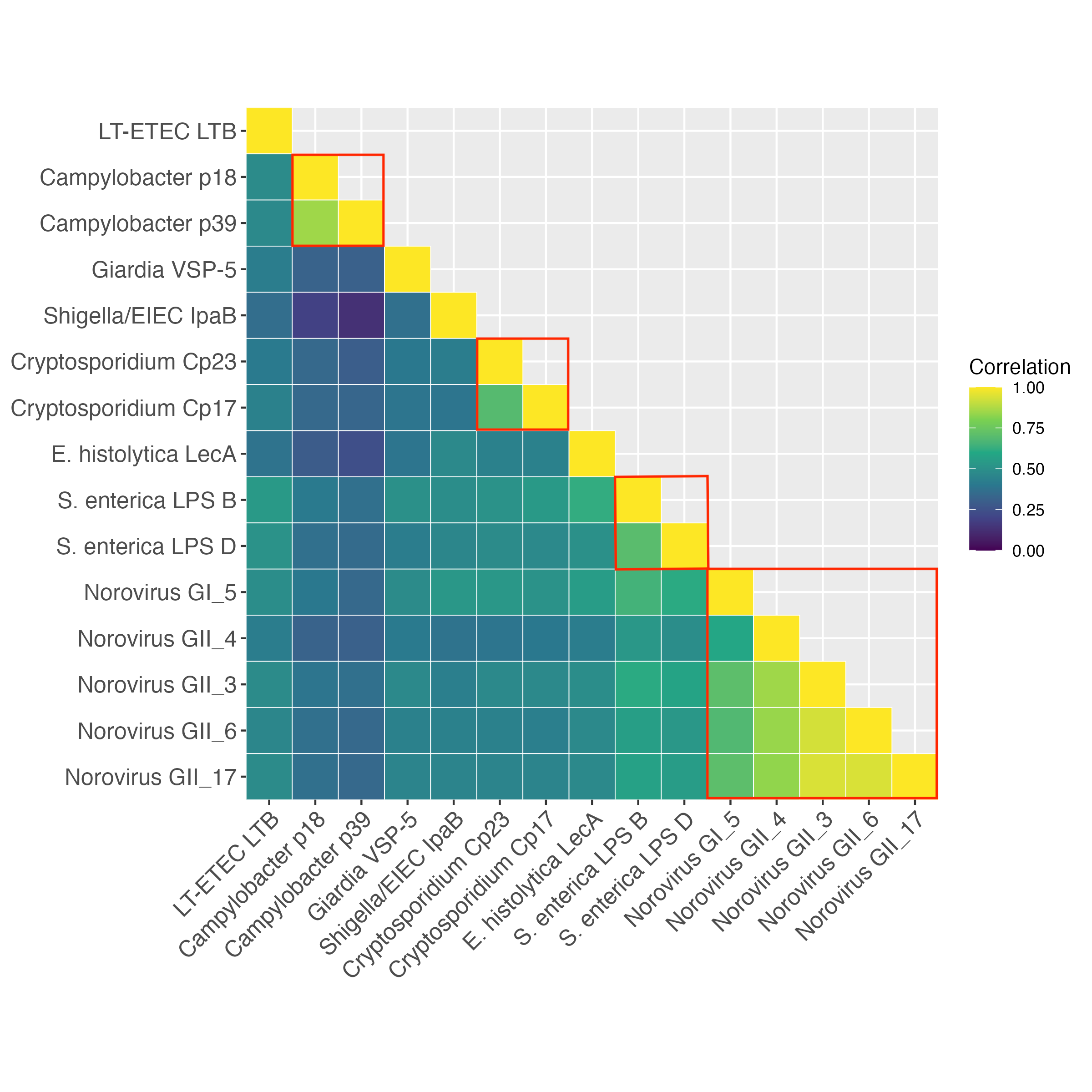

**Fig. S1. Sample-level correlation between IgG serology responses to enteric pathogen antigens.** Correlation coefficients were calculated using Pearson correlation on log10-transformed Luminex Median Fluorescence Intensity minus background (MFI-bg) values for individual antigens at the sample level (n = 1575 measurements). Antigens are ordered according to hierarchical clustering (average linkage, distance = 1 − Pearson correlation). Red boxes highlight antigens from the same pathogen.

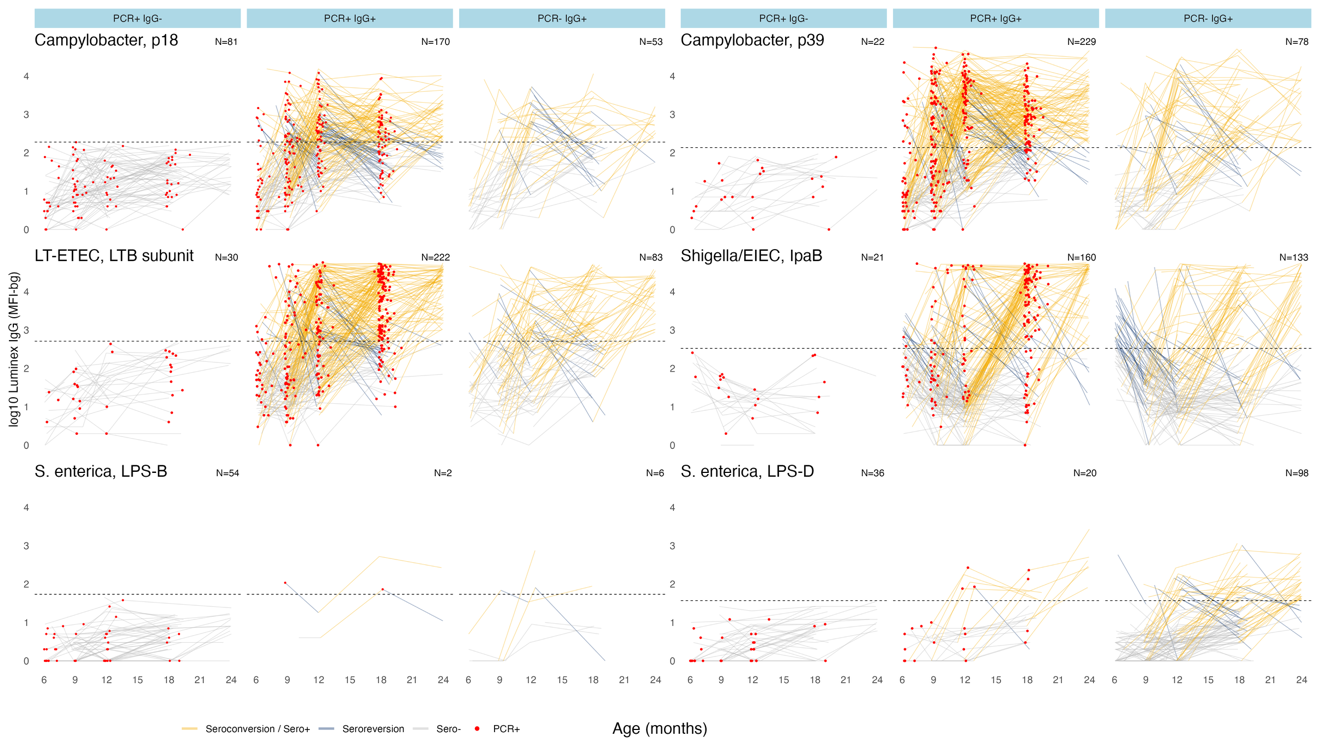

**Fig. S2.** **IgG mean fluorescent intensity (MFI) of bacterial antigens in children over the study period.** Log10(MFI) is represented for each child-set that had a positive PCR result and a negative IgG result (IgG-, PCR+), a negative PCR result and a positive IgG result (PCR- IgG+), or positive PCR and positive IgG (PCR+ IgG+) over the course of the study. PCR positive results were only considered if the result was at any timepoint before the last recorded IgG result to account for seroconversion time. Red points represent PCR+ results that were taken at the same timepoint an IgG result (IgG+ or IgG-) was available. If there was no IgG result at that timepoint (NA), the point was shifted to next corresponding IgG result in the sequence for that child. Lines are colored according to whether the child seroconverted or remained seropositive (yellow), seroreverted (blue), or remained seronegative (grey) according to the seropositivity threshold for that antigen (dashed line).

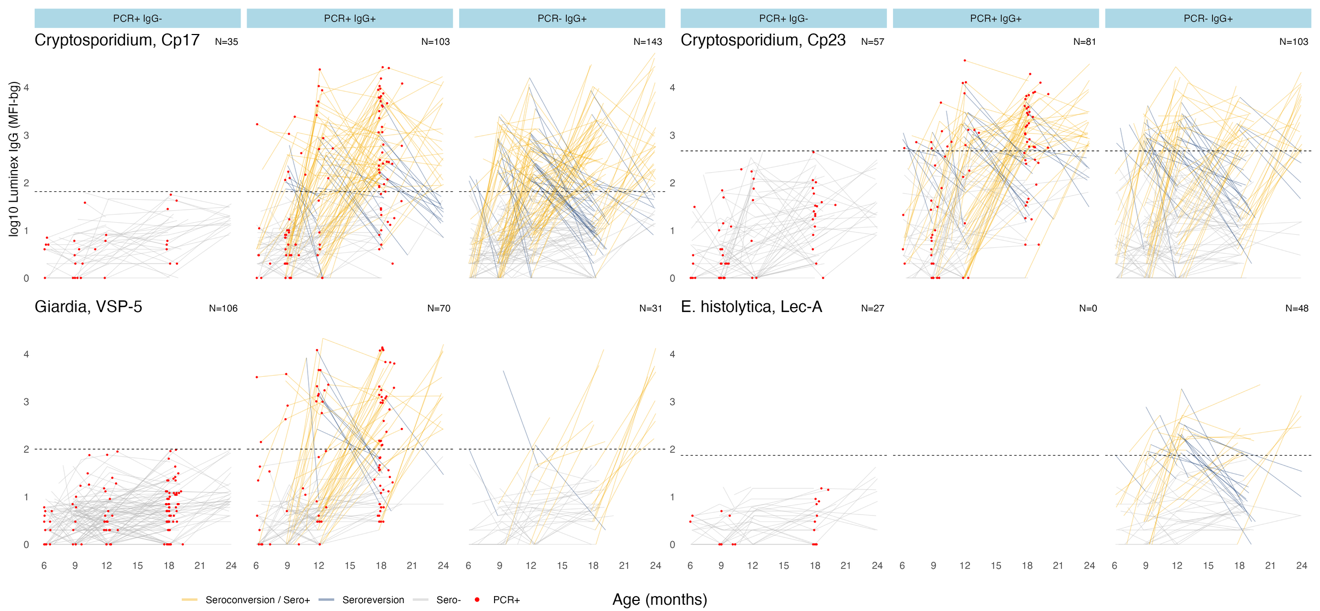

**Fig S3. IgG mean fluorescent intensity (MFI) of protozoal antigens in children over the study period.** Log10(MFI) is represented for each child-set that had a positive PCR result and a negative IgG result (PCR+ IgG-), a negative PCR result and a positive IgG result (PCR- IgG+), or positive PCR and positive IgG (PCR+ IgG+) over the course of the study. PCR positive results were only considered if the result was at any timepoint before the last recorded IgG result to account for seroconversion time. Red points represent PCR+ results that were taken at the same timepoint an IgG result (IgG+ or IgG-) was available. If there was no IgG result at that timepoint (NA), the point was shifted to next corresponding IgG result in the sequence for that child. Lines are colored according to whether the child seroconverted or remained seropositive (yellow), seroreverted (blue), or remained seronegative (grey) according to the seropositivity threshold for that antigen (dashed line).

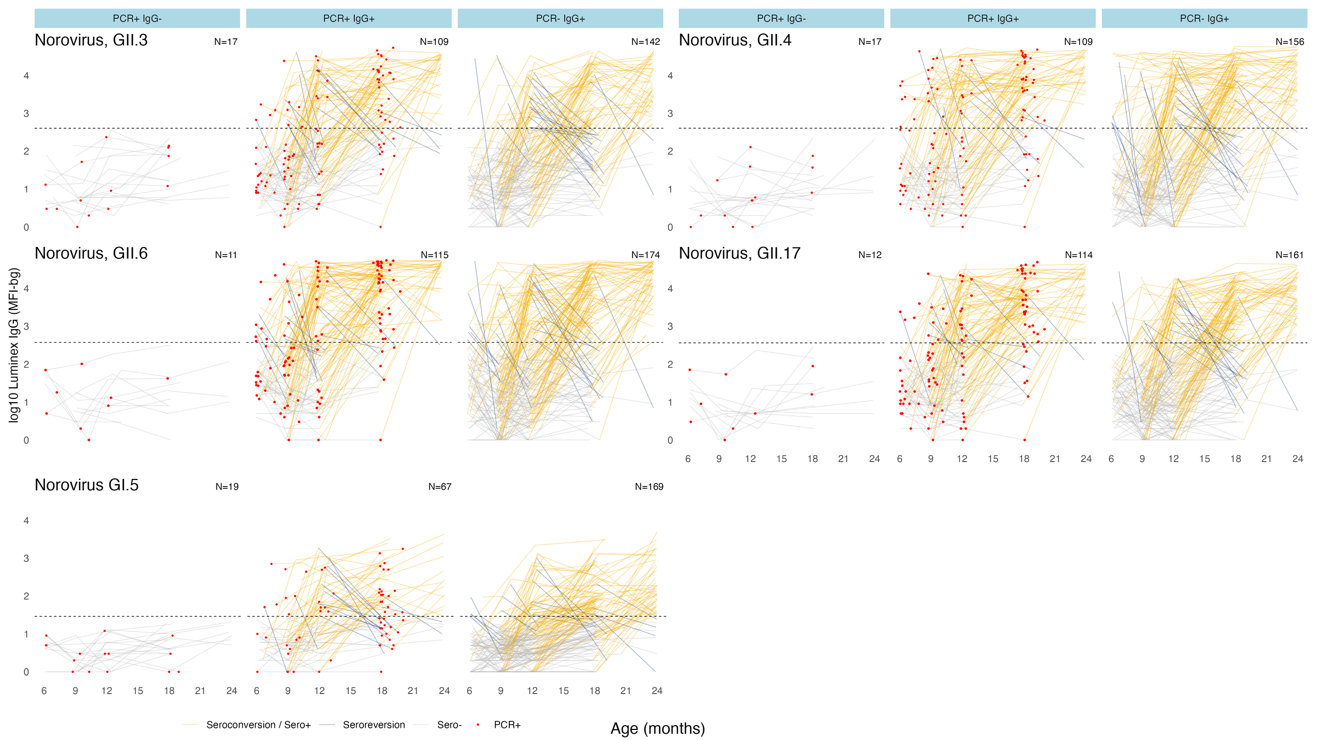
**Fig S4. IgG mean fluorescent intensity (MFI) of viral antigens in children over the study period.** Log10(MFI) is represented for each child-set that had a positive PCR result and a negative IgG result (PCR+ IgG-), a negative PCR result and a positive IgG result (PCR- IgG+), or positive PCR and positive IgG (PCR+ IgG+) over the course of the study. PCR positive results were only considered if the result was at any timepoint before the last recorded IgG result to account for seroconversion time. Red points represent PCR+ results that were taken at the same timepoint an IgG result (IgG+ or IgG-) was available. If there was no IgG result at that timepoint (NA), the point was shifted to next corresponding IgG result in the sequence for that child. Lines are colored according to whether the child seroconverted or remained seropositive (yellow), seroreverted (blue), or remained seronegative (grey) according to the seropositivity threshold for that antigen (dashed line).

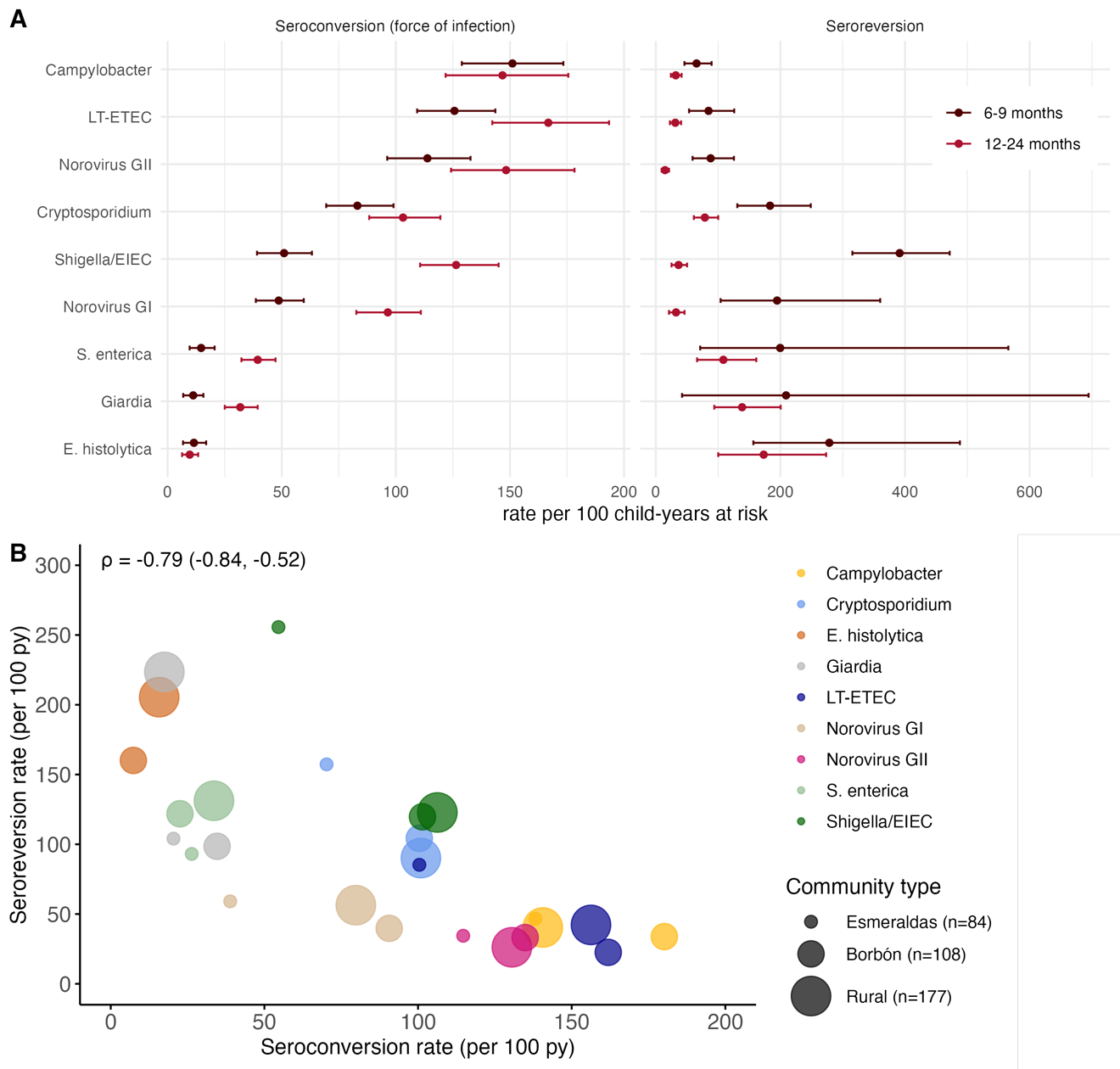
**Fig S5. Age specific seroconversion and seroreversion rates among 369 children measured longitudinally in coastal Ecuador.** We estimated nonparametric longitudinal rates of seroconversion and seroreversion among children at risk from ages 6-9 months and 12-24 months, assigning each event to the midpoint of the corresponding measurement interval (A). Error bars represent 95% confidence intervals as estimated via nonparametric bootstrap, resampling children with replacement to account for within-child correlation. IgG response measured in multiplex using median fluorescence units minus background (MFI-bg) on the Luminex platform (N = 1,575 measurements from 369 children). We calculated spearman correlation coefficients (ρ) directly from the observed pathogen-community estimates (B), 95% confidence intervals for ρ were estimated using nonparametric bootstrap, resampling children with replacement. One point was excluded to optimize visualization (Esmeraldas; *Entamoeba histolytica*, SCR: 2.96 per 100 pys; SRR 785 per 100 pys).

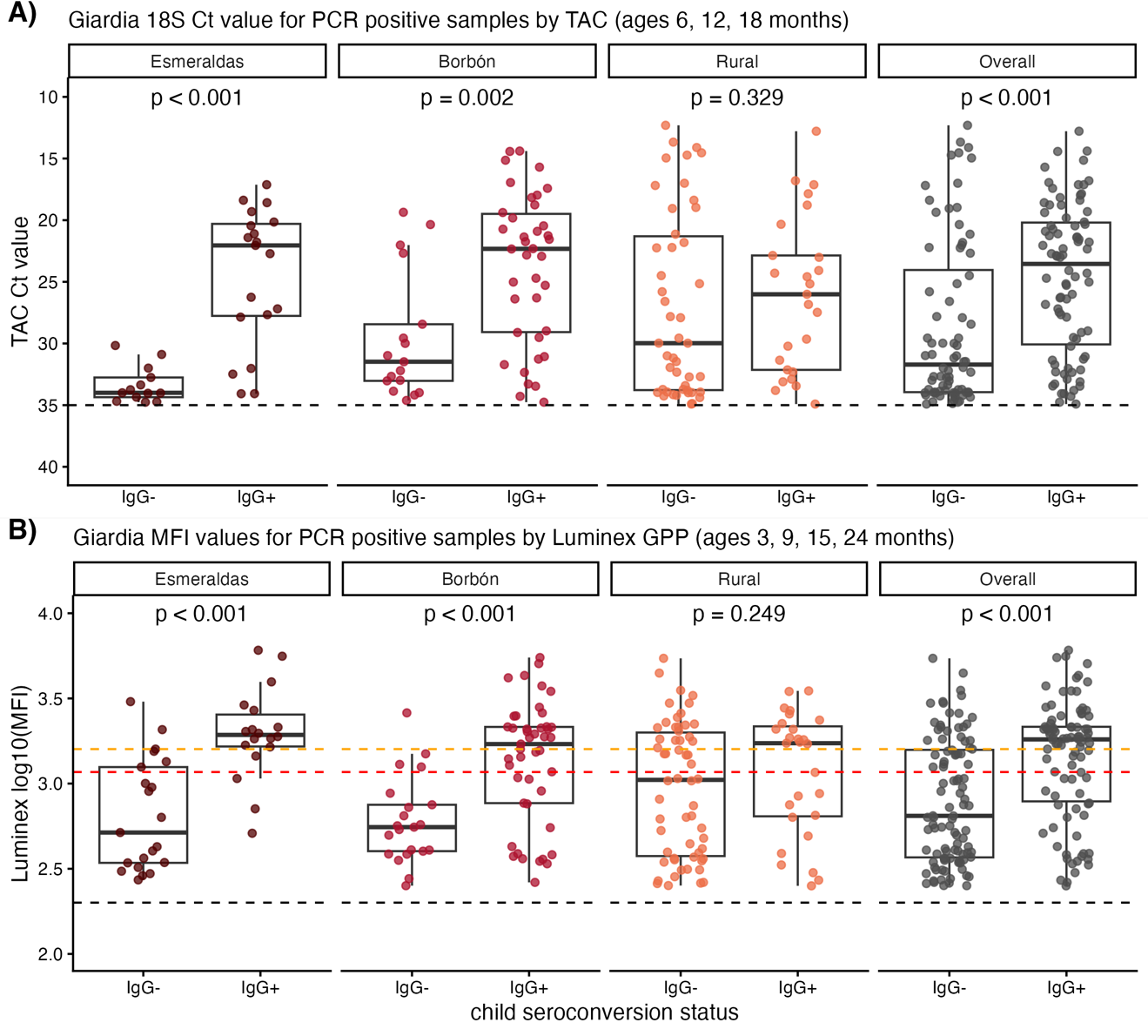
**Fig S6.** **PCR assay values for children who tested positive for Giardia by PCR, grouped by seroconversion status over the study period.** Ct values were measured using the TaqMan PCR (TAC) assay at 6, 12, and 18 months, and log10-transformed MFI values were measured using the Luminex xTAG-GPP assay at 3, 9, 15, and 24 months. PCR+ values that occurred at or after the last recorded IgG result and were the only PCR+ result for that child set, were excluded to account for seroconversion time. Horizontal black dashed lines indicate assay-specific thresholds for PCR positivity. Red dashed line indicates low positive and limit of detection and yellow dashed line indicates moderate positive results, as reported by xTAG GPP manufacturer (10). Mann–Whitney U tests showed significant differences between seroconversion groups for both assays for all samples (TaqMan: p-value = 4.74 × 10⁻⁶; Luminex: p-value = 0.00264). This difference was less pronounced for rural communities compared to Borbón and the city of Esmeraldas.

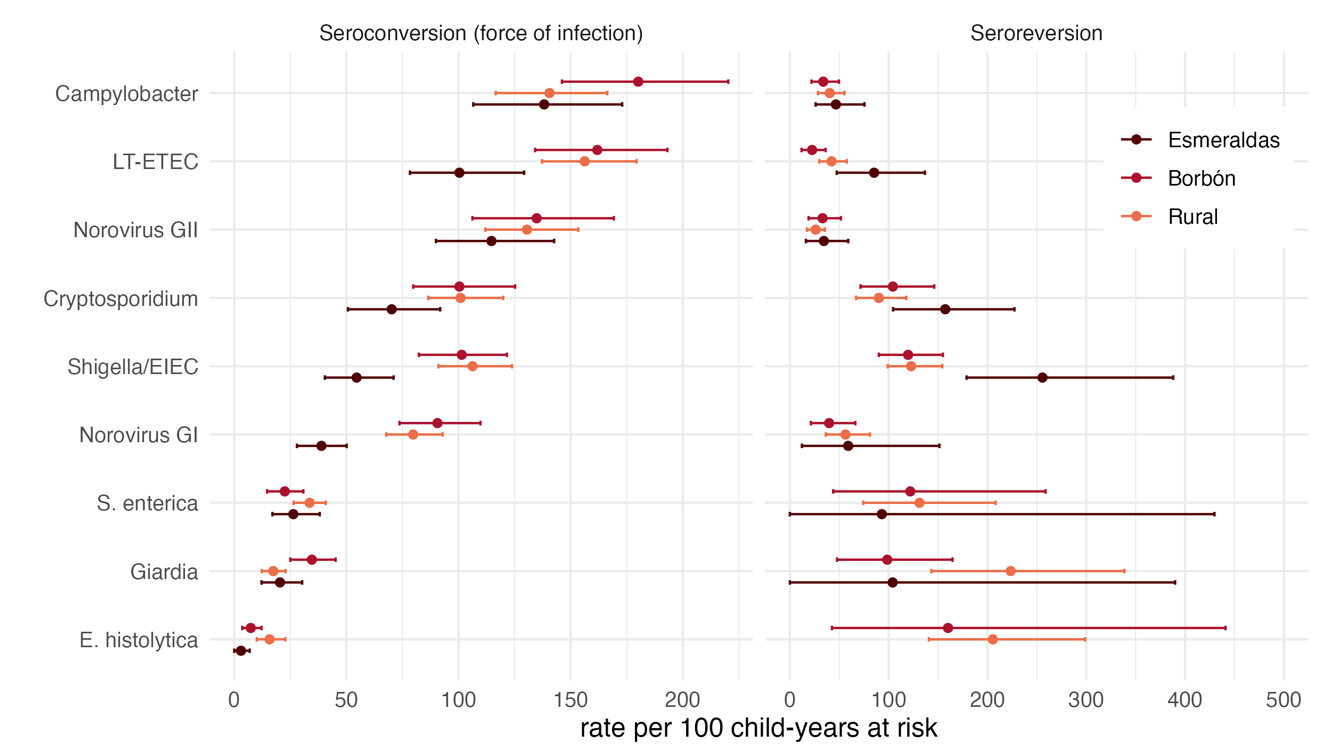

**Fig S7. Community specific seroconversion and seroreversion rates among 369 children aged 6 to 24 months measured longitudinally in coastal Ecuador.** We estimated nonparametric longitudinal rates of seroconversion and seroreversion among children at risk, assigning each event to the midpoint of the corresponding measurement interval. Error bars represent 95% confidence intervals as estimated with bootstrapping, resampling children with replacement to account for within-child correlation. IgG response measured in multiplex using median fluorescence units minus background (MFI-bg) on the Luminex platform (N = 1575 measurements from 369 children). Due to small number of children who seroreverted to *E. histolytica* (n = 3), we could not estimate the seroreversion rate for the city of Esmeraldas.

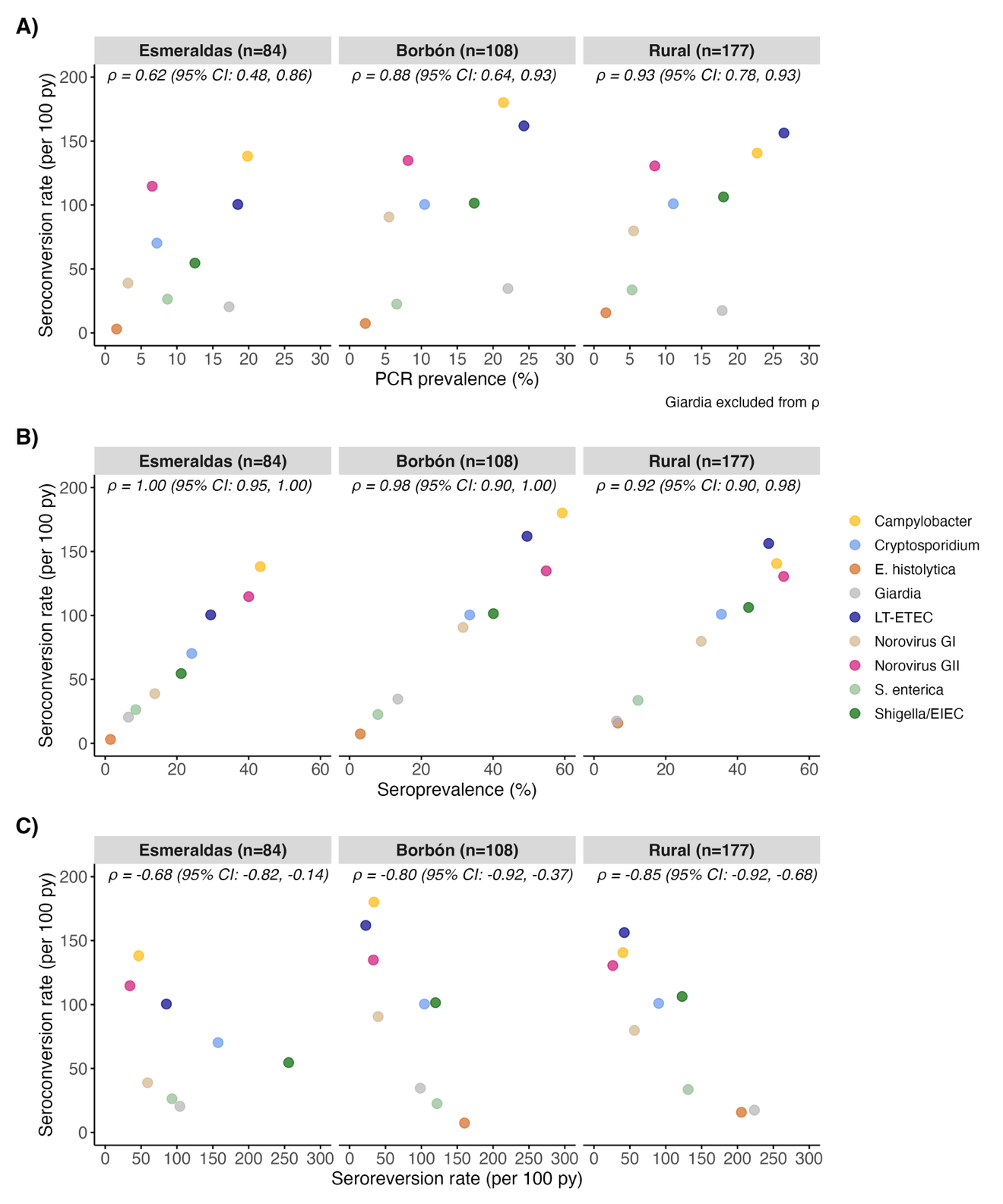

**Fig S8. Relationship between measures of infection across pathogens by community type.** Seroconversion rates and seroprevalence were estimated over five visits from ages 6 to 24 months (n=369 total children, 1575 total samples). PCR-detected prevalence was estimated over seven visits from ages 3 to 24 months (n=369 total children, 2229 total samples). 95% confidence intervals for Spearman correlation estimates (ρ) were estimated using a nonparametric bootstrap, resampling children with replacement separately within each community (Esmeraldas, n = 85; Borbón, n = 108; Rural, n = 177). **A.** IgG seroconversion rates compared with PCR detected prevalence in stool. *Giardia* was excluded from spearman rho (ρ) calculation because the relationship between measures differs markedly from other pathogens in this setting (details in Results). With *Giardia* included the spearman correlation (ρ) is as follows city of Esmeraldas, ρ=0.52, 95^th^ CI: 0.25-0.68; Borbón, ρ=0.62, 95^th^ CI: 0.42-0.78; Rural, ρ=0.75, 95^th^ CI: 0.52-0.77). **B.** Incident seroconversion rates compared with seroprevalence across all visits. **C.** Incident seroconversion rates compared with incident seroreversion rates across all visits.

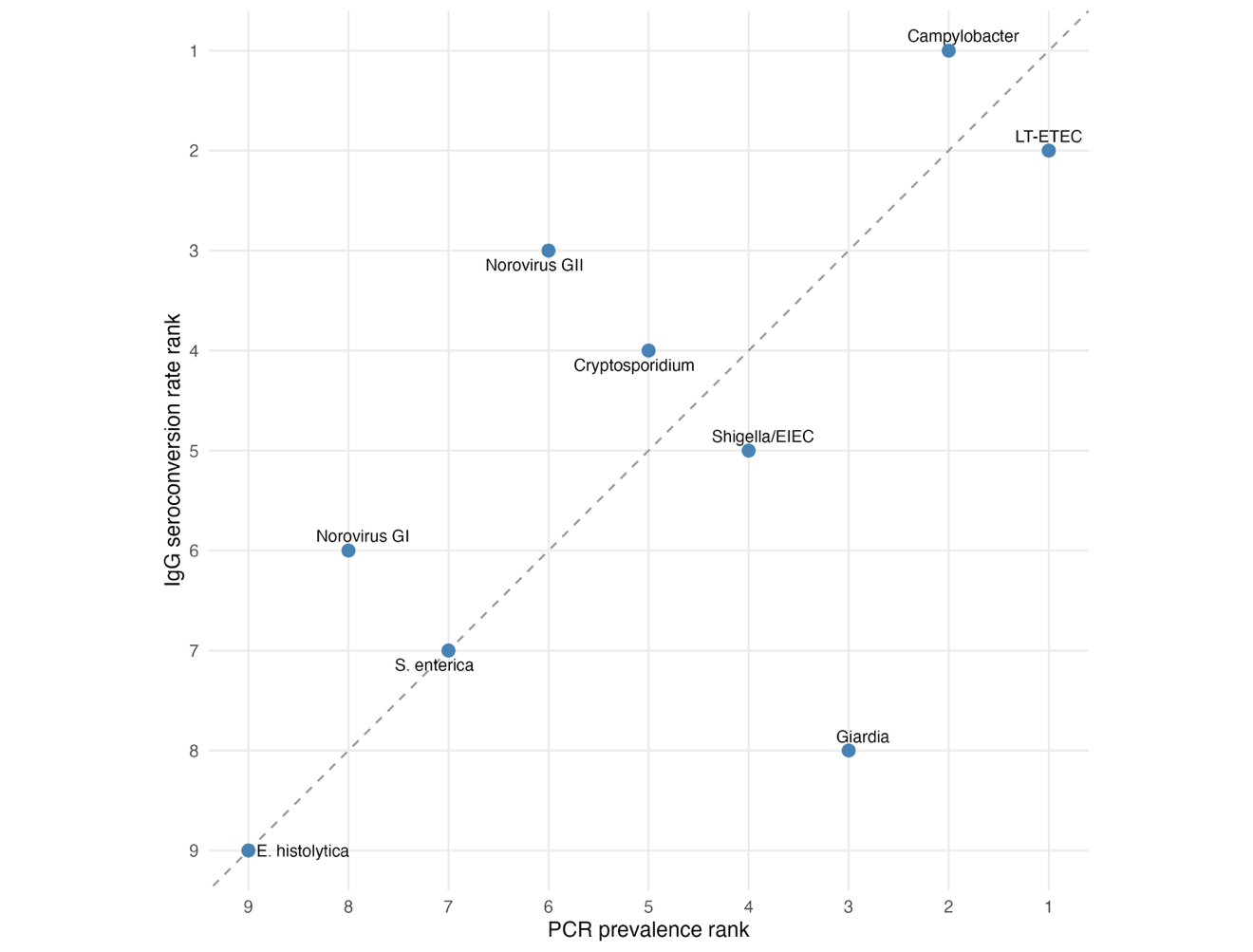

**Fig S9:** **Relative ranking of PCR detected prevalence and IgG seroconversion rate of pathogens.** Each point represents one of the nine pathogens assayed by both platforms, ranked from lowest (1) to highest (9) by PCR detected prevalence over the study periods or IgG seroconversion rate within each assay. The dashed line indicates perfect rank concordance. PCR detected prevalence and IgG seroconversion rate were calculated from all samples tested by each assay (IgG, n=369 total children, 1575 total samples; PCR, n=369 total children, 2229 total samples).

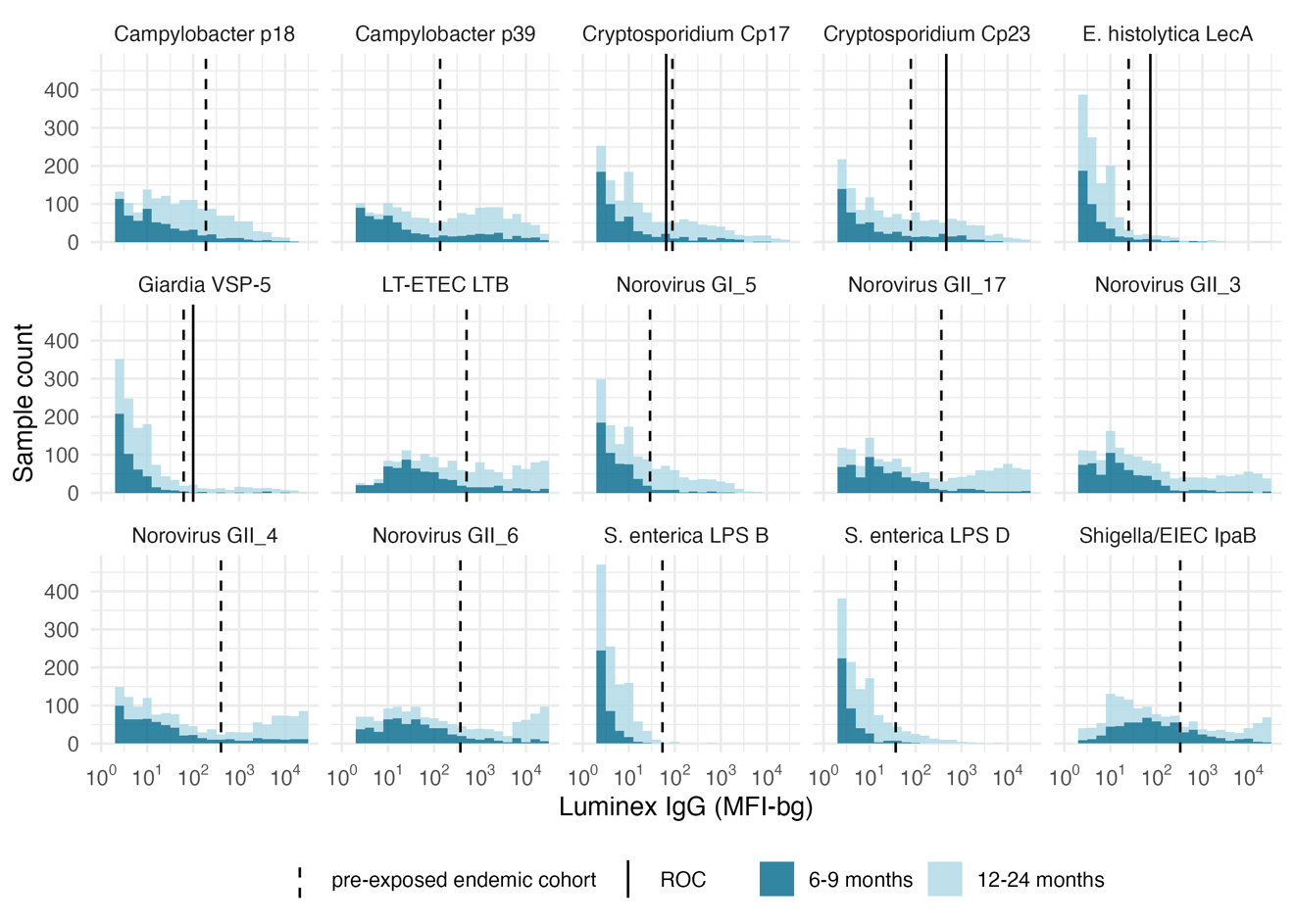
**Fig S10.** **IgG distributions among children and seropositivity cutoffs for enteric pathogen antigens included in the multiplex bead assay.** Sample distributions are highlighted for the 6-9-month and the 12–24-month age ranges to better-illustrate separation between seronegative and seropositive values before IgG boosting and waning at older ages. Seropositivity thresholds were determined using receiver operator characteristic (ROC) curve analysis of known positive and negative samples (solid line) or through a pre-exposed endemic cohort approach (dashed line). ROC-based cutoffs were used if available. For pathogens with multiple antigens, samples with IgG above the seropositivity threshold for any antigen were considered seropositive.

Tables

Table S1. Number of blood and stool samples (369 children) tested by month of age. Luminex xTAG-GPP assay was used for 3, 9, 15 and 24 month stool sample testing. A custom TaqMan array card assay was used for 6, 12, 18 month stool sample testing.

|  | **Months (N)** | | | | | | |  |
| --- | --- | --- | --- | --- | --- | --- | --- | --- |
|  | **3** | **6** | **9** | **12** | **15** | **18** | **24** | **Total samples (N)** |
| **Stool (PCR)** | 339 | 339 | 337 | 329 | 297 | 297 | 291 | 2,229 |
| **Blood (IgG)** | 0 | 342 | 327 | 312 | 0 | 304 | 290 | 1,575 |

Table S2. Characteristics of study children that contributed dried blood spot samples, by community type

|  | **Esmeraldas** | **Borbón** | **Rural Road** | **Rural River** | **Overall** |
| --- | --- | --- | --- | --- | --- |
|  | **(N=84)** | **(N=108)** | **(N=137)** | **(N=40)** | **(N=369)** |
| **Sex** |  |  |  |  |  |
| Female | 35 (41.7%) | 53 (49.1%) | 73 (53.3%) | 16 (40.0%) | 177 (48.0%) |
| **Age of Mother (years)** |  |  |  |  |  |
| Mean (SD) | 26.6 (6.03) | 25.3 (5.59) | 26.4 (5.05) | 28.5 (6.50) | 26.4 (5.66) |
| **Mother Ethnicity*** |  |  |  |  |  |
| Afro-Ecuadorian | 44 (52.4%) | 67 (62.0%) | 121 (88.3%) | 39 (97.5%) | 271 (73.4%) |
| Mestizo | 39 (46.4%) | 33 (30.6%) | 12 (8.8%) | 1 (2.5%) | 85 (23.0%) |
| Manabi | 1 (1.2%) | 5 (4.6%) | 3 (2.2%) | 0 (0%) | 9 (2.4%) |
| Chachi | 0 (0%) | 2 (1.9%) | 0 (0%) | 0 (0%) | 2 (0.5%) |
| **Mother’s education*** |  |  |  |  |  |
| Primary or less | 2 (2.4%) | 10 (9.3%) | 16 (11.7%) | 10 (25.0%) | 38 (10.3%) |
| Secondary or less | 68 (81.0%) | 67 (62.0%) | 93 (67.9%) | 28 (70.0%) | 256 (69.4%) |
| Post-secondary or greater | 14 (16.7%) | 31 (28.7%) | 27 (19.7%) | 2 (5.0%) | 74 (20.1%) |
| **Child birth order*** |  |  |  |  |  |
| 1 | 22 (26.2%) | 36 (33.3%) | 30 (21.9%) | 9 (22.5%) | 97 (26.3%) |
| 2 | 32 (38.1%) | 33 (30.6%) | 33 (24.1%) | 10 (25.0%) | 108 (29.3%) |
| 3+ | 30 (35.7%) | 39 (36.1%) | 73 (53.3%) | 21 (52.5%) | 163 (44.2%) |
| **Wealth quartile** |  |  |  |  |  |
| 1 (poorest) | 18 (21.4%) | 36 (33.3%) | 33 (24.1%) | 8 (20.0%) | 95 (25.7%) |
| 2 | 18 (21.4%) | 27 (25.0%) | 31 (22.6%) | 16 (40.0%) | 92 (24.9%) |
| 3 | 18 (21.4%) | 14 (13.0%) | 47 (34.3%) | 11 (27.5%) | 90 (24.4%) |
| 4 (wealthiest) | 30 (35.7%) | 31 (28.7%) | 26 (19.0%) | 5 (12.5%) | 92 (24.9%) |
| **Household crowding** |  |  |  |  |  |
| ≥ 3 people per room | 18 (21.4%) | 21 (19.4%) | 14 (10.2%) | 1 (2.5%) | 54 (14.6%) |
| **Sanitation*** |  |  |  |  |  |
| Basic | 84 (100%) | 91 (84.3%) | 123 (89.8%) | 23 (57.5%) | 321 (87.0%) |
| Limited | 0 (0%) | 7 (6.5%) | 9 (6.6%) | 7 (17.5%) | 23 (6.2%) |
| Unimproved | 0 (0%) | 8 (7.4%) | 2 (1.5%) | 10 (25.0%) | 20 (5.4%) |
| Open defecation | 0 (0%) | 1 (0.9%) | 2 (1.5%) | 0 (0%) | 3 (0.8%) |
| **Water Source *** |  |  |  |  |  |
| Basic | 81 (96.4%) | 103 (95.4%) | 133 (97.1%) | 40 (100%) | 357 (96.7%) |
| Limited/Unimproved | 0 (0%) | 3 (2.8%) | 1 (0.7%) | 0 (0%) | 4 (1.1%) |
| Surface water | 0 (0%) | 2 (1.9%) | 1 (0.7%) | 0 (0%) | 3 (0.8%) |
| ***E. coli* in water source** | |  |  |  |  |
| Detected* | 36 (42.9%) | 66 (61.1%) | 102 (74.5%) | 31 (77.5%) | 235 (63.7%) |
| **Flooring*** |  |  |  |  |  |
| Cement/finished | 73 (86.9%) | 52 (48.1%) | 108 (78.8%) | 11 (27.5%) | 244 (66.1%) |
| Earth/sand/palm/bamboo | 5 (6.0%) | 5 (4.6%) | 2 (1.5%) | 1 (2.5%) | 13 (3.5%) |
| Wooden boards | 6 (7.1%) | 50 (46.3%) | 27 (19.7%) | 28 (70.0%) | 111 (30.1%) |

**Data categorized as “missing” or “other” were not included in table*

**Table S3. Overall Pathogen Prevalence. Prevalence was calculated by dividing the total number of positive samples by the total number of samples collected over the course of the study.**

| **Pathogen** | **Assay** | **Positives (N)** | **Total samples (N)** | **Prevalence (Positives / Total samples; (%, 95^th^ CI)** |
| --- | --- | --- | --- | --- |
| *Campylobacter* | PCR | 484 | 2229 | 21.7 (20.1, 23.5) |
|  | IgG | 815 | 1575 | 51.7 (48.6, 54.8) |
| *Cryptosporidium*^†^ | PCR | 211 | 2112 | 10 (8.8, 11.4) |
|  | IgG | 511 | 1575 | 32.4 (29.8, 35.2) |
| *Entamoeba* *histolytica*^†^ | PCR | 40 | 2229 | 1.8 (1.3, 2.5) |
|  | IgG | 70 | 1574 | 4.4 (3.3, 6) |
| *Giardia* | PCR | 422 | 2229 | 18.9 (17.2, 20.8) |
|  | IgG | 133 | 1575 | 8.4 (7, 10.2) |
| LT-ETEC^†^ | PCR | 535 | 2225 | 24 (22.3, 25.9) |
|  | IgG | 705 | 1575 | 44.8 (42, 47.5) |
| Norovirus GI^†^ | PCR | 111 | 2229 | 5 (4.1, 6) |
|  | IgG | 424 | 1574 | 26.9 (24.5, 29.5) |
| Norovirus GII | PCR | 177 | 2229 | 7.9 (6.9, 9.2) |
|  | IgG | 798 | 1575 | 50.7 (47.4, 53.9) |
| *Salmonella enterica^*^* | PCR | 62 | 965 | 6.4 (5.1, 8.1) |
|  | IgG | 160 | 1575 | 10.2 (8.6, 12) |
| *Shigella*/EIEC | PCR | 370 | 2229 | 16.6 (15.2, 18.1) |
|  | IgG | 590 | 1575 | 37.5 (34.8, 40.2) |

**Salmonella enterica* PCR results were only available from the TaqMan array assay

^†^Small subset of samples did not pass quality control for this antigen or PCR target

**Table S4. Rank order of pathogen PCR detected prevalence and IgG seroconversion rate, by community type. PCR detected prevalence and IgG seroconversion rate were calculated from all samples tested by each assay over the study period (IgG, n=369 total children, 1575 total samples; PCR, n=369 total children, 2229 total samples).**

| **Community type** | **Pathogen** | **PCR prevalence** | **PCR prevalence rank** | **IgG SCR (per 100 py)** | **IgG SCR rank** | **Rank difference** |
| --- | --- | --- | --- | --- | --- | --- |
| Esmeraldas | *Campylobacter* | 19.8% (16.6–23.5%) | 1 | 138.2 (106.6–173.0) | 1 | 0 |
| Esmeraldas | LT-ETEC | 18.5% (15.6–21.8%) | 2 | 100.4 (78.4–129.3) | 3 | 1 |
| Esmeraldas | *Giardia* | 17.3% (13.8–21.3%) | 3 | 20.4 (12.2–30.3) | 8 | 5 |
| Esmeraldas | *Shigella*/EIEC | 12.5% (9.9–15.6%) | 4 | 54.6 (40.4–71.1) | 5 | 1 |
| Esmeraldas | *S. enterica* | 8.7% (5.8–12.8%) | 5 | 26.3 (17.0–38.1) | 7 | 2 |
| Esmeraldas | *Cryptosporidium* | 7.2% (5.2–9.8%) | 6 | 70.2 (50.7–91.8) | 4 | -2 |
| Esmeraldas | Norovirus GII | 6.5% (4.7–9.0%) | 7 | 114.7 (90.0–142.6) | 2 | -5 |
| Esmeraldas | Norovirus GI | 3.2% (1.9–5.2%) | 8 | 38.9 (28.0–50.2) | 6 | -2 |
| Esmeraldas | *E. histolytica* | 1.6% (0.8–3.1%) | 9 | 3.0 (0.0–6.9) | 9 | 0 |
| Borbon | LT-ETEC | 24.3% (21.2–27.7%) | 1 | 161.9 (134.2–193.1) | 2 | 1 |
| Borbon | *Giardia* | 22.1% (18.3–26.4%) | 2 | 34.6 (25.0–45.2) | 7 | 5 |
| Borbon | *Campylobacter* | 21.4% (18.5–24.7%) | 3 | 180.1 (146.1–220.3) | 1 | -2 |
| Borbon | *Shigella*/EIEC | 17.4% (14.6–20.6%) | 4 | 101.4 (82.3–121.6) | 4 | 0 |
| Borbon | *Cryptosporidium* | 10.5% (8.3–13.1%) | 5 | 100.4 (79.8–125.2) | 5 | 0 |
| Borbon | Norovirus GII | 8.1% (6.2–10.6%) | 6 | 134.8 (106.2–169.3) | 3 | -3 |
| Borbon | *S. enterica* | 6.6% (4.2–10.2%) | 7 | 22.5 (14.7–30.8) | 8 | 1 |
| Borbon | Norovirus GI | 5.5% (3.8–7.7%) | 8 | 90.6 (73.7–109.8) | 6 | -2 |
| Borbon | *E. histolytica* | 2.2% (1.2–3.8%) | 9 | 7.4 (3.6–12.2) | 9 | 0 |
| Rural | LT-ETEC | 26.5% (23.8–29.3%) | 1 | 156.3 (137.2–179.4) | 1 | 0 |
| Rural | *Campylobacter* | 22.7% (20.4–25.3%) | 2 | 140.6 (116.6–166.3) | 2 | 0 |
| Rural | *Shigella/E*IEC | 18.0% (16.1–20.1%) | 3 | 106.3 (91.1–123.8) | 4 | 1 |
| Rural | *Giardia* | 17.9% (15.7–20.3%) | 4 | 17.4 (12.3–22.9) | 8 | 4 |
| Rural | *Cryptosporidium* | 11.1% (9.2–13.2%) | 5 | 100.9 (86.5–120.0) | 5 | 0 |
| Rural | Norovirus GII | 8.5% (6.9–10.3%) | 6 | 130.5 (112.0–153.4) | 3 | -3 |
| Rural | Norovirus GI | 5.5% (4.3–7.0%) | 7 | 79.8 (67.8–93.0) | 6 | -1 |
| Rural | *S. enterica* | 5.3% (3.6–7.8%) | 8 | 33.6 (26.5–40.8) | 7 | -1 |
| Rural | *E. histolytica* | 1.7% (1.0–2.7%) | 9 | 15.8 (10.1–22.8) | 9 | 0 |

**Table S5. Enteric pathogen targets use in multiplex serology assay, adapted from (11).**

| **Reagent type or resource** | **Designation** | **Source or reference** | **Identifiers** | **Additional information** |
| --- | --- | --- | --- | --- |
| Peptide, recombinant protein | *Giardia intestinalis*  VSP-5 | PMID: 11500396 PMID: 20876825 | GenBank: HM036223 | Dr. Jeffrey Priest (CDC) |
| Peptide, recombinant protein | *Cryptosporidium* Cp17 | PMID: 10699255 PMID: 15165066 | GenBank: AF114166 | Dr. Jeffrey Priest (CDC) |
| Peptide, recombinant protein | *Cryptosporidium* Cp23 | PMID: 8892291 PMID: 10203492 | GenBank: U34390 | Dr. Jeffrey Priest (CDC) |
| Peptide, recombinant protein | *Entamoeba histolytica*, LecA | PMID: 2000392 PMID: 14741152 PMID: 24591430 | GenBank: M60498 | Dr. William Petri (University of Virginia) and Dr. Joel Herbein (TechLab) |
| Peptide, recombinant protein | *Campylobacter jejuni p18* | PMID: 8576327 PMID: 15754430 | GenBank: X83374 | Dr. Jeffrey Priest (CDC) |
| Peptide, recombinant protein | *Campylobacter jejuni* p39 | PMID: 10688204 PMID: 15754430 | GenBank: CAL34198.1 | Dr. Jeffrey Priest (CDC) |
| Peptide, recombinant protein | ETEC heat labile toxin B (LTB) subunit | PMID: 3882744 PMID: 18494692 |  | Sigma-Aldrich |
| Native antigen | *Salmonella enterica* LPS group B | PMID: 2567429 PMID: 17329442 |  | Sigma-Aldrich |
| Native antigen | *Salmonella enterica* LPS group D | PMID: 2567429 PMID: 17329442 |  | Sigma-Aldrich |
| Peptide, recombinant protein | *Shigella/EIEC*, IpaB | PMID: 23583496 | Genbank: AF330458 | Dr. Edward T. Ryan (Massachusetts General Hospital and WRAIR ) |
| Virus like particles | Norovirus GI.5 | This paper |  | Dr. Jan Vinje (CDC) |
| Virus like particles | Norovirus GII.3 | PMID: 31896578 | AB365435 | Dr. Robert Atmar (Baylor College of Medicine)  Gretchen Cooley (CDC) |
| Virus like particles | Norovirus GII.4 | PMID: **31424386** |  | Dr. Jan Vinje (CDC) |
| Virus like particles | Norovirus GII.6 | PMID: 25428247 | GU930737 | Dr. Robert Atmar (Baylor College of Medicine)  Gretchen Cooley (CDC) |
| Virus like particles | Norovirus GII.17 | PMID: 25428247 | DQ438972 | Dr. Robert Atmar Baylor College of Medicine)  Gretchen Cooley (CDC) |

**Table S6. Enteric pathogen targets use in multiplex serology assay, adapted from (11)**

| **Enteric pathogen** | **PCR Target** | **Primer sequences** |
| --- | --- | --- |
| ***Cryptosporidium*** | 18S | F, GGGTTGTATTTATTAGATAAAGAACCA  R, AGGCCAATACCCTACCGTCT  P, FAM-TGACATATCATTCAAGTTTCTGAC-MGB |
| ***Campylobacter*** | cadF (*jejuni/coli*)^*^ | F, CWGCTAAACCATARAAATAAAATTTCTCAC  R, YTTTGAAGGTAATTTAGATATGGATAATCG  P, VIC-CATTTTGAYGATTTTTGGCTTGA-MGB |
|  | GlyA (*coli)^*^* | F, AAACCAAAGCTTATCGTGTGC  R, AGTGCAGCAATGTGTGCAAT  P, FAM-TAAGCTCCAACTTCATCCG-MGB |
|  | hipO (*jejuni)^*^* | F, CTTGCGGTCATGATGGACATAC  R, AGCACCACCCAAACCCTCTTCA  P, FAM-TGCTTGCTGCAAAGTATT-MGB |
| *Entamoeba histolytica* | 18S | F, ATTGTCGTGGCATCCTAACTCA  R, GCGGACGGCTCATTATAACA  P, FAM-TCATTGAATGAATTGGCCATTT-MGB |
| *Giardia* | 18S | F, GACGGCTCAGGACAACGGTT  R, TTGCCAGCGGTGTCCG  P, FAM-CCCGCGGCGGTCCCTGCTAG-MGB |
| Norovirus GI | ORF1-ORF2 | F, CGYTGGATGCGNTTYCATGA  R, CTTAGACGCCATCATCATTYAC  P, FAM-TGGACAGGAGATCGC-MGB |
| Norovirus GII | ORF1-ORF2 | F, CARGARBCNATGTTYAGR TGGATGAG  R, TCGACGCCATCTTCATTCACA  P, FAM-TGGGAGGGCGATCGCAATCT-MGB |
| LT-ETEC | LT | F, TTCCCACCGGATCACCAA  R, CAACCTTGTGGTGCATGATGA  P, FAM-CTTGGAGAGAAGAACCCT-MGB |
| *Salmonella enterica* | ttr | F, CTCACCAGGAGATTACAACATGG  R, AGCTCAGACCAAAAGTGACCATC  P, FAM-CACCGACGGCGAGACCGACTTT-MGB |
| *Shigella/EIEC* | ipaH | F, CCTTTTCCGCGTTCCTTGA  R, CGGAATCCGGAGGTATTGC  P, VIC-CGCCTTTCCGATACCGTCTCTGCA-MGB |
|  | virF | F, TCTGAGGAGGAGGTTTCTATCGATT  R, GAAACAGCTGATAAAAGGCAAGCT  P, FAM-CCGAAAGGCATCTCTTTT-MGB |
